# Behavioural factors influencing hand hygiene practices across domestic, institutional, and public community settings: A systematic review

**DOI:** 10.1101/2025.03.11.25323561

**Authors:** Bethany A. Caruso, Jedidiah S. Snyder, Lilly A. O’Brien, Erin LaFon, Kennedy Files, Dewan Muhammad Shoaib, Sridevi K. Prasad, Hannah Rogers, Oliver Cumming, Joanna Esteves Mills, Bruce Gordon, Marlene K. Wolfe, Matthew C. Freeman

**Affiliations:** Hubert Department of Global Health, Rollins School of Public Health, Emory University, Atlanta, GA, USA; Gangarosa Department of Environmental Health, Rollins School of Public Health, Emory University, Atlanta, GA, USA; Woodruff Health Sciences Center Library, Emory University, Atlanta, GA, USA; Department of Disease Control, London School of Hygiene and Tropical Medicine, London, UK; Water, Sanitation, Hygiene and Health Unit, World Health Organization, Geneva, Switzerland

## Abstract

This systematic review sought to understand barriers and enablers to hand hygiene in community settings. Eligible studies addressed hand hygiene in a community setting, included a qualitative component, and were published in English between January 1, 1980, and March 29, 2023. Studies were excluded if in healthcare settings or were animal research. We searched PubMed, Web of Science, EMBASE, CINAHL, Global Health, Cochrane Library, Global Index Medicus, Scopus, PAIS Index, WHO IRIS, UN Digital Library and World Bank eLibrary, manually searched relevant systematic reviews’ reference lists, and consulted experts. We used MaxQDA Software to code papers, using the COM-B framework to classify barriers and enablers. We used thematic analysis to describe each COM-B sub-theme identified, GRADE-CerQual to assess confidence in evidence for thematic findings, and the Mixed Method Appraisal Tool (MMAT) to assess risk of study bias. Eighty studies were included; most took place in Africa (31; 39%), South-East Asia (31; 39%) and domestic settings (54; 68%). The mean MMAT score was 4.86 (good quality). Barriers and/or enablers were reported across all COM-B constructs and sub-constructs. The most reported barriers aligned with Physical Opportunity (e.g., soap availability), Reflective Motivation (e.g., hand hygiene not prioritized) and Automatic Motivation (e.g., no habit). In contrast, the most reported enablers aligned with Automatic (i.e., habit) and Reflective (i.e., perception of health risk) Motivation. Findings confirm that lacking necessary resources for hand hygiene hinders practice, even when people are motivated. Results may explain why hand hygiene increases when there are acute health risks (e.g., COVID), but decreases when risks are perceived to fade. The qualitative methodology used among the studies may have revealed a broader array of barriers and enablers than what might have been found by quantitative, researcher-driven studies, but representativeness may be limited. Findings can inform the design of future hand hygiene initiatives.

**What is already known on this topic:** Hand hygiene prevents disease, but barriers like limited access to soap, water, and competing priorities hinder practice. Most reviews focus narrowly on specific behaviours, like handwashing with soap and water exclusively, rather than a broader suite of hand hygiene behaviours, or on specific contexts, like schools, rather than a broader range of community settings.

**What this study adds:** This study systematically identifies barriers and enablers to hand hygiene across various community settings using an established behavioral framework. It highlights the importance of contextual and behavioral factors, showing that resource provision is essential, but alone may not instigate or sustain hand hygiene practices without addressing broader motivational and habitual drivers.

**How this study might affect research practice or policy:** Findings from this study can guide the design of targeted interventions that not only ensure the availability of resources but also foster habits and address motivational barriers to hand hygiene. Policymakers can leverage these insights to develop more comprehensive hand hygiene programs, while researchers can explore under-investigated areas such as the barriers and drivers to hand hygiene in community settings among people with disabilities.

## 1. INTRODUCTION

Hand hygiene, whether through handwashing with water and soap or other methods such as the use of alcohol-based hand rubs (ABHR), is a critical public health measure. Hand hygiene interventions are relatively inexpensive to implement ^1^ and can prevent several infectious diseases, including enteric ^2^ and respiratory ^3^ infections, which account for a large burden of disease ^4^ and high healthcare costs ^5^. Establishing global guidelines and recommendations is essential to guide hand hygiene initiatives, protect public health, and strengthen resilient health systems ^6^.

According to the Ottawa Charter, community settings are where ‘health is created and lived by people within the setting of their everyday life; where they learn, work, play, and love,’ ^7^ and include domestic, public, and institutional spaces ^8^. Guidelines for hand hygiene in healthcare settings are well-established^9–12^, and additional guidelines emphasize investing in hand hygiene as a core public health measure ^13–16^.

Despite the recognized importance of hand hygiene, and the unprecedented prioritization driven by the COVID-19 pandemic ^16^, gaps and inconsistencies remain in global guidance on specific measures ^17^. A recent scoping review identified 51 existing international guidelines and highlighted a lack of consistent evidence-based recommendations and identified four areas where clear recommendations are needed for hand hygiene in community settings: (1) effective hand hygiene; (2) minimum requirements; (3) behavior change; and (4) government measures ^8^.In their review, MacLeod et al. 2023 found that guidelines within the scoping review provided inconsistent recommendations on determinants of hand hygiene to target for interventions and behavior change models or frameworks. Understanding what determinants hinder or enable hand hygiene behavior is necessary for designing appropriate interventions and for assessing if existing interventions appropriately respond to the barriers and enablers identified. Existing systematic reviews on determinants of hand hygiene have been informative but limited in scope. Two recent reviews only included studies that focused on handwashing with soap and water ^18,19^. Other syntheses have been narrowed in eligible settings including studies with settings likely to include children (e.g., schools, homes) ^20^, hand hygiene practices among primary and secondary school students in sub-Saharan Africa ^21^, and prevalence and determinants of hand hygiene behavior in India ^22^. There remains a need for a more comprehensive review to understand the suite of barriers and enablers to a broader range of hand hygiene practices across community settings.

This review seeks to understand the suite of barriers and enablers to hand hygiene in community settings. The research question for this review was identified through an extensive consultation process by the WHO with external experts ^23,24^, following a scoping review of current international guidelines ^8^. Findings from the study can inform intervention design in varied settings to enable improved hand hygiene and reduced transmission of infectious diseases within community settings.

## 2. METHODS

### 2.1. Research questions

This systematic review primarily sought to assess the following question: what are key behavioral barriers and enablers to practicing effective hand hygiene in community settings?

In addition, we aimed to understand which theories, models, and frameworks have been used in studies investigating barriers and enablers of hang hygiene in community settings and which have been the most common.

### 2.2. Search strategy

This review was pre-registered with PROSPERO (registration number: CRD42023429145) and is reported in accordance with the Preferred Reporting Items for Systematic Reviews and Meta-Analyses ^25^ (PRISMA) criteria (See S1 – PRISMA Checklist). This review was part of an integrated protocol for multiple related reviews to synthesize the evidence for effective hand hygiene in community settings ^24^. We adopted a two-phased approach for identifying relevant studies. Phase 1 involved a broad search to capture all studies on hand hygiene in community settings that were relevant across multiple related systematic reviews. The outcome of phase 1 was a reduced sample from which further screening, specific to this review, was performed. A full description of the procedures followed for searches, study inclusion, outcomes data collection, analysis, and reporting of the multiple related reviews is presented in the published protocol ^24^.

This search included studies published between January 1, 1980, and March 29, 2023, and published in English—unless the title and abstract was published in English and/or a non-English language article was referenced in an existing systematic review. We searched 12 peer-reviewed and grey literature databases. PubMed, Web of Science, EMBASE (Elsevier), CINAHL (EBSCOhost), Global Health (CAB), Cochrane Library, Global Index Medicus, Scopus (Elsevier), Public Affairs Information Service (PAIS) Index (ProQuest) were searched on March 23, 2023 and WHO Institutional Repository for Information Sharing (IRIS), UN Digital Library, and World Bank eLibrary were searched on March 28, 2023 using search terms related to hand hygiene broadly and restrictions on terms related to healthcare settings in the titles. We searched trial registries (International Clinical Trials Registry Platform, clinicaltrials.gov) for trials related to hand hygiene in community settings on March 29, 2023.

We conducted manual searches of reference lists of two relevant reviews ^18,19^. For reviews that provided a list of the reviewed articles, we searched only those references. If a list was not available, we searched all references and screened for potentially relevant titles. These reviews had 113 total references of which 51 were duplicates, 33 were already identified in our database search, and 29 were added to phase 2 title and abstract screening. We contacted 35 content experts and organizations, using snowballing methods, between April to May 2023 for information on relevant unpublished literature.

### 2.3. Selection criteria

Studies were eligible for inclusion if they addressed hand hygiene in community settings, included a qualitative component, were in English, and published between January 1, 1980, and March 29, 2023. For this review, hand hygiene refers to any hand cleansing undertaken for the purpose of removing or deactivating pathogens from hands and efficacious hand hygiene is defined as any practice which effectively removes or deactivates pathogens from hands and thereby has the potential to limit disease transmission ^10^. The term community settings included domestic (e.g., households), public (e.g., markets, public transportation hubs, vulnerable populations [e.g., people experiencing homelessness], parks, squares, or other public outdoor spaces, shops, restaurants, and cafes), and institutional (e.g., workplace, schools and universities, places of worship, prisons and places of detention, nursing homes and long-term care facilities) spaces ^8^. Studies were excluded if they were in healthcare settings or were animal research. Studies in nursing homes and long-term care facilities were excluded as part of phase 2 screening as these were determined to be similar to evidence generated in healthcare settings. There were no geographic restrictions.

Studies that include a qualitative component (i.e., qualitative and mixed methods studies) were eligible for inclusion in the analysis. Studies that include qualitative research are best suited to address the research question as they leverage open-ended questions to understand phenomena, perspectives, and lived experiences as voiced by participants themselves. In contrast, any barriers or enablers identified in quantitative research will have been pre-identified for investigation based on their inclusion in a survey by the research team. As a result, the survey itself limits the suite of barriers and enablers identified. To report on the scope of research on barriers and enablers of hand hygiene in community settings— regardless of study design—we identified the library of purely quantitative studies focused on enablers and barriers to practicing effective hand hygiene in community settings, which we provide as a supplement.

We used Covidence software for systematic reviews ^26^. In both phases, screening of each article (phase 1 – title and abstract only; phase 2 – title and abstract, then full text review) was performed independently by two reviewers, with discordance between reviewers reconciled by a third review. The stages and related outcomes of the search and screening process are described in the PRISMA flow chart (S2- Figure 1).

**Figure 1.**
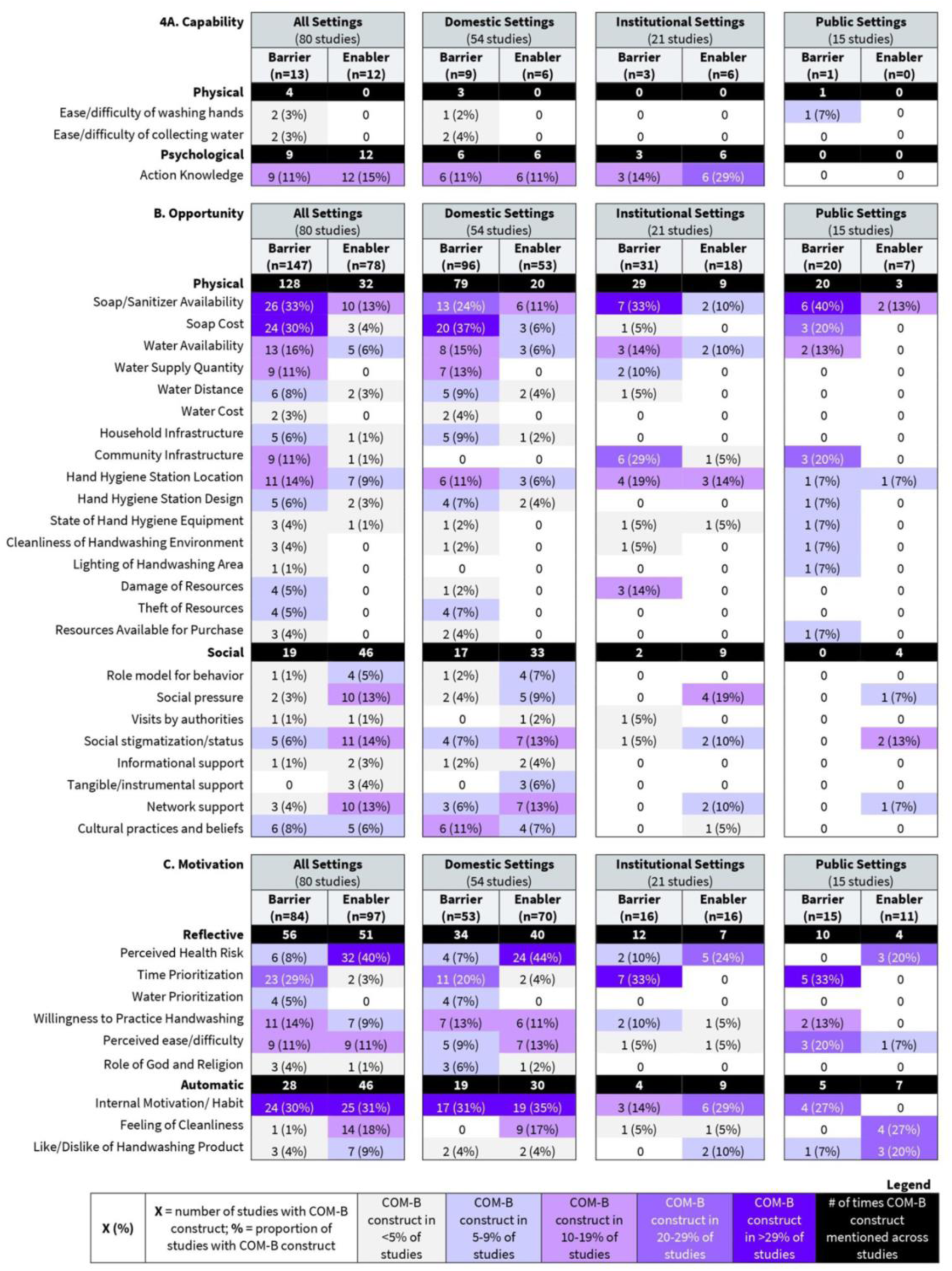
Frequency of participant-reported barriers and enablers to hand hygiene practices in community settings categorized by (A) Capability, (B) Opportunity, and (C) Motivation components of the COM-B framework.

### 2.4. Data extraction and analysis

Reviewers independently double extracted data from each included study using a customized data extraction tool and assessed risk of bias for each article using the Mixed Method Appraisal Tool (MMAT) ^27,28^. Any conflicts between reviewers over bias assessment were resolved by discussion. All data extraction tools are provided in the supplement (S3 – Covidence Extraction Questionnaire; S4 – Data Extraction Spreadsheet)

For appraising the quality of mixed methods studies using the MMAT, the individual study components were assessed using the appropriate categories: the qualitative component, the quantitative category for the quantitative component, and the mixed-methods category. Final scores, which can range from 0-5 across study types (5 is the best), are presented in the main text of the results only for qualitative components of the MMAT assessment because only qualitative data were extracted from the mixed methods studies included in the review. We provide our grading for both the qualitative and mixed methods components in the supplement.

To provide a broad overview of included studies, we extracted information about study characteristics, including the region, location, setting, participants, and hand hygiene practice explored.

To understand which theories, models, and frameworks have been leveraged in research investigating barriers and enablers of hand hygiene in community settings and which have been the most common, we extracted the names of any theories, models, or frameworks authors reported to have used (if any). We did not extract data on or report how theory was used (e.g., in tool design and/or analysis).

To classify the types of barriers and enablers of hand hygiene practices in community settings reported in and across studies and settings, we used the COM-B framework ^29^ as a guide. COM-B has been used to categorize barriers and enablers in multiple systematic reviews ^30–32^, including hand hygiene ^20^. The COM-B model proposes that three components, Capability, Opportunity, and Motivation, interact to enable and maintain behavior. Capability can be psychological (knowledge) or physical (skills); opportunity can be social (societal influences) or physical (environmental resources); motivation can be automatic (emotion) or reflective (beliefs, intentions). Table 1 provides comprehensive definitions of each COM-B construct and examples related to hand hygiene.

**Table 1.** Definitions and examples of constructs from the COM-B framework used to code barriers and enablers to practice effective hand hygiene.

| COM-B construct | Definition<sup>1</sup> | Examples |
| --- | --- | --- |
| **Capability** | **Knowledge, skills and abilities required to engage in a particular behavior** |  |
| **Physical Capability** | Physical strength, skill or stamina | Physical ability to wash hands |
| **Psychological Capability** | Knowledge/ psychological strength, skills or stamina | Knowledge on how to wash hands |
| **Opportunity** | **External factors which make the execution of a particular behavior possible** |  |
| **Physical Opportunity** | Opportunities provided by the environment, such as time, location and resource | Availability of soap |
| **Social Opportunity** | Opportunities as a result of social factors, like cultural norms and social cues | Cultural norm to wash hands |
| **Motivation** | **The internal processes which influence decision making and behaviors** |  |
| **Automatic Motivation** | Automatic processes, such as our desires, impulses and inhibitions | Have habit of washing hands |
| **Reflective Motivation** | Reflective processes, such as making plans and evaluating things that have already happened | Believe handwashing reduces illness risk |

<sup>1</sup>Definitions from (Social Change UK)

To identify barriers and enablers in relevant qualitative data, included studies were imported into MAXQDA (version 12) ^33^ qualitative analysis software. Following best practice, ^34^ line-by-line coding of the results sections, which present the empirical data from the studies, was carried out twice by independent team members. The team used a code book developed *a priori to* categorize barriers and enablers to practicing hand hygiene in community settings into the six sub-components of the COM-B framework (Table 1). The codebook was pre-reviewed by one of the creators of the COM-B framework and is provided in the supplement (S5 – COM-B Codebook). Text coded in MAXQDA may have included quotes directly from primary study participants (first-order data) and/or text from authors summarizing or synthesizing information from their study participants (second-order data).

To assist in aggregating and synthesizing findings from the line-by-line coding in MAXQDA, reviewers simultaneously used a customized data extraction tool in excel (S4 - Data Extraction Spreadsheet) to note if a particular COM-B-related barrier or enabler was identified in each study. Each time a particular barrier or enabler was identified and coded in a study using MAXQDA, the research team member also noted that the specific barrier or enabler was identified on the customized excel data extraction sheet and cut and pasted the relevant coded text as a means of supporting the coding decision. A third team member not involved in the line-by-line coding then reviewed all codes and copied text segments in the excel file to confirm that codes were accurately applied given the copied supporting text.

We identified the frequency with which COM-B-categorized barriers and enablers were identified *across* studies and report these frequencies overall and by setting. We do not report the frequency of coded COM-B-categorized barriers and enablers identified *within* studies because some studies may simply provide more depth on or give additional mention of the same barriers or enablers at different points in the paper based on their analysis strategies. Frequencies, therefore, were only used to report on the extent to which barriers or enablers were identified across studies.

To provide depth on the identified barriers and enablers of hand hygiene practices in community settings categorized using COM-B, we used a thematic analysis approach ^35^ to describe each COM-B sub-theme identified by setting type and if a barrier or enabler. We used all coded text related to the sub-theme for this analysis, enabling multiple codes from the same study to inform the summary. For example, we examined all codes on the theme of ‘Physical Opportunity: Soap Availability’ to summarize insights from the qualitatively coded data by setting type (domestic, institutional, public) and if perceived to hinder or enable handwashing. Importantly, for both thematic analysis and tabulation numerically, we aimed to only include the coded text as a barrier or enabler if a distinct connection was made between the sub-theme and hand hygiene behavior. In many instances, studies reported on resources available or absent in a particular setting (e.g., soap availability at school), but if the specific text did not report a specific connection between soap availability and whether and how it influenced hand hygiene, it was not included.

To assess the level of confidence in evidence for each thematic finding, we used the GRADE-CerQual approach ^36^. Specifically, for each identified barrier or enablers in a subtheme, we assessed the methodological limitations, coherence, adequacy, and relevance by setting; each criterion was characterized as follows: ‘no or very minor concerns’, ‘minor concerns’, ‘moderate concerns’, ‘serious concerns’. The methodological limitations criteria assessment was informed by the MMAT scores. We leveraged information from the studies involved in each sub-theme for the remaining criteria. Specifically, we looked across the coded text that informed the sub-theme to assess coherence (fit between primary study data and the synthesized review finding) question, assessed the number of studies informing the theme to assess adequacy, and reviewed the location, setting, and populations engaged to in the aligned studies to assess relevance ^37–40^. All criteria were used to make a final assessment of confidence for each sub-theme (high, moderate, low, very low) ^41^.

### Ethics and Patient/ Public Involvement Statement

Because we reviewed published documents, no ethical approval was required. Patients or the public were not involved directly in the design, or conduct, or reporting, or dissemination plans of our research. This evidence synthesis supports the forthcoming WHO Guidelines for Hand Hygiene in Community Settings; the study questions were developed in broad consultation with a network of key partners. Findings from this review will be disseminated alongside the Guidelines.

### Data Availability

All data and extraction templates will be made publicly available upon publication.

## 3. RESULTS

### 3.1. Characteristics of the studies included in this review

We identified 80 studies that met our inclusion criteria (S2-PRISMA Flow Diagram and S6-Overview of Included Studies). Studies represent all WHO regions, with almost 80% from Africa (31; 39%) and South-East Asia (31; 39%) (Table 2). Most studies focused on domestic settings (54; 68%), followed by institutional (21; 26%) and public (15; 19%) settings (accounting for studies focused on more than one setting). Within institutional settings, schools were the most studied (14; 67%), with fewer studies in other institutional settings such as workplaces (4; 19%), universities (2; 10%), or childcare centers (1; 5%). Within public settings, internally displaced person’s camps were most common, (7; 47%), followed by markets (5; 33%) and refugee camps (1; 7%). The largest proportion of studies (24; 30%) focused on both men and women for handwashing behavior; 23% (18) focused on children (boys and girls), 10% (8) targeted women specifically, 7% (5) engaged food workers, and 35% (28) focused on the general population or did not specify. Handwashing with soap and water was the most common specified hand hygiene practice of focus (60; 75%); 18% (14) of studies did not provide detail about the specific hand hygiene practice of focus. Because only two (2.5%) studies, focused on alcohol-based hand rubs, we refer to ‘handwashing’ as the behavior throughout the results. Regarding specific contexts or ‘risk scenarios’ within which the studies took place, 15% (12) were related to COVID-19 and 10% (8) focused on internal displacement. A total of 987 studies comprise the quantitative library, which is included in the online dataset ( [to be made public upon publication]).

**Table 2.** Characteristics of the included studies.

| **Descriptive characteristics of studies** | **Total**<br>**n (%)** |
| --- | --- |
| **Total number of studies** | **80 (100.0)** |
| **Region\*** |  |
| African Region | 31 (38.8) |
| South-East Asian Region | 31 (38.8) |
| European Region | 7 (8.8) |
| Region of the Americas | 7 (8.8) |
| Eastern Mediterranean Region | 3 (3.8) |
| Western Pacific Region | 2 (2.5) |
| **Location\*** |  |
| Rural | 35 (43.8) |
| Urban | 23 (28.8) |
| Peri-urban | 5 (6.3) |
| Unspecified | 29 (36.3) |
| **Setting\*** |  |
| Domestic (Households) | 54 (67.5) |
| Institutional | 21 (26.3) |
| Schools | 14 (66.6) |
| Workplaces | 4 (19.0) |
| Universities | 2 (9.5) |
| Childcare centers | 1 (4.7) |
| Public | 15 (18.8) |
| Internally displaced persons’ camps | 7 (46.6) |
| Refugee camps | 1 (6.6) |
| Markets | 5 (33.3) |
| Other | 2 (13.3) |
| **Study participants\*** |  |
| Adults – Women and men | 24 (30.0) |
| Children – Girls and boys | 18 (22.5) |
| Adults – Women | 8 (10.0) |
| Food workers | 5 (6.3) |
| Other | 5 (6.3) |
| General population/unspecified | 28 (35.0) |
| **Specific hand hygiene practice explored\*** |  |
| Handwashing with soap and water | 60 (75.0) |
| Handwashing with water only | 3 (3.8) |
| Handwashing with soap alternatives | 1 (1.3) |
| Hand rubbing with alcohol-based rub | 2 (2.5) |
| Handwashing - unspecified | 14 (17.5) |
| **Risk Scenario** |  |
| COVID-19 | 12 (15.0) |
| Internal displacement | 8 (10.0) |
| Flu | 1 (1.3) |
| Typhoid Fever | 2 (2.5) |
| HIV | 1 (1.3) |
| Landslide disaster | 1 (1.3) |
| People who inject drugs | 1 (1.3) |
| General emergency setting | 1 (1.3) |
| **Study quality rating (mean)\*\*** | **4.86** |

**Note:** \*Descriptive statistics for region, location, setting, primary outcome studied, hand hygiene practice, and hand hygiene intervention represent variables that were multi-select. \*\*Quality appraisal using the Mixed Method Appraisal Tool (Pluye and Hong 2014; Hong et al. 2018): Possible scores are 0–5 across study types (5 is the best).

### 3.2. Quality of the studies included in this review

The mean study quality was 4.86 overall indicating good quality (5 is the best). The quality appraisal scores for each study are in the supplement (S7 – MMAT Assessment).

### 3.3. Theory, model, or framework use in included studies

A third of the included studies reported use of a theory, model or framework (26; 33%) (Table 3). The earliest study included in our review that reported using theory was published in 1994 (S8 – Theory Bar chart). Thirteen different theories, models, or frameworks were reported to be used; those most commonly used included *IBM-WASH (Integrated Behavioural Model for Water, Sanitation, and Hygiene)* (8; 31%) ^42^, *Behaviour Centered Design/Evo-Eco Model* (7; 27%) ^43^, *Health Belief Model* (4; 15%) ^44^, and *COM-B (Capability, Opportunity, Motivation, and Behavior)* (4; 15%) ^29^.

**Table 3.** Theories or models reported to be used in included studies. (N=80)

| Theory | Total<br>n (%) |
| --- | --- |
| Study did not report using theory | 54 (67.0) |

| Study did report using theory* | 26 (33.0) |
| --- | --- |
| IBM-WASH | 8 (30.8) |
| Behaviour Centered Design/Evo-Eco Model | 7 (26.9) |
| Health Belief Model | 4 (15.4) |
| COM-B | 4 (15.4) |
| RANAS | 2 (7.7) |
| Theory of Planned Behavior | 2 (7.7) |
| Theoretical Domains Framework | 2 (7.7) |
| Elicitation Procedure | 1 (3.8) |
| Cultural Consensus Theory | 1 (3.8) |
| Prototype Theory | 1 (3.8) |
| Donabedian's Model for Assessing Quality of Care | 1 (3.8) |
| PRECEDE-PROCEED Model | 1 (3.8) |
| The Model of Families Use of Health Service | 1 (3.8) |

**Note:** \*Multiple theories could have been reported per study so the sum of theories used (35) is greater than the total number of studies that report using theory (26).

### 3.4. COM-B-categorized barriers and enablers of hand hygiene practices across community settings

Across all settings, barriers and/or enablers were reported across all COM-B constructs and sub-constructs, though some constructs were more frequently reported as barriers while others were more frequently reported as enablers (Figure 1). Specifically, the most reported barriers overall and across settings aligned with *Physical Opportunities* (i.e., ‘soap availability’, ‘cost of soap’, ‘water availability’), *Reflective* Motivation (i.e., ‘time prioritization’) and *Automatic Motivation* (i.e., ‘internal motivation/habit’). Several sub-themes of water—related to *Physical Opportunity*—also were noted as barriers for different reasons and with varied frequency. If assessed in combination, water also emerges as a substantial barrier (‘water availability’ (13/80, 16%); ‘water supply quantity’ (9/80, 11%); ‘water distance’ (6/80, 7%); all water collectively (28/80, 35%)).

In contrast, the most reported enablers aligned with *Reflective Motivation (i.e., ‘disease risk’)* and *Automatic Motivation* (i.e., ‘internal motivation/habit’).

### 3.5. COM-B-categorized barriers and enablers of hand hygiene practices in domestic settings

In domestic settings, the most reported barriers were related to *Physical Opportunity* (‘cost of soap’ (20/54; 36%) and ‘soap availability’ (13/54; 24%)), *Automatic Motivation* (‘internal motivation/habit’ (17/54; 31%)) and *Reflective* Motivation (‘time prioritization’ (11/54; 20%). Further, if assessed together, water-related barriers were a commonly noted barrier in domestic settings (‘water availability’ (8/54, 15%); ‘water supply quantity’ (7/54, 13%); ‘water distance’ (5/54, 9%); all water collectively (20/54, 37%)).

Based on thematic analysis (S9- Thematic Analysis Table), studies found that the lack of soap available in domestic settings and the cost of soap (both *Physical Opportunity)* hindered handwashing practices; confidence in these themes was high (S10- CERQual Assessment). Regarding water-related themes, studies described both a lack of water and irregular water supply as hindrances. Distance to water sources hindered handwashing altogether or required people to limit/ration the amount of water people used for handwashing, if practiced (confidence in these water themes was moderate and high).

Regrading ‘internal motivation/habit’ (*Automatic Motivation;* high confidence), study participants noted that they lacked a routine or habit for practicing hand hygiene; they specifically said they would forget or not wash hands due to ‘laziness’ or noted that they have other habits or routines already established that do not include handwashing, thus hindering practice. Related to ‘time prioritization’ (*Reflective Motivation;* high confidence), studies discussed that the amount of time required for handwashing prevented them from the behavior altogether or handwashing with soap specifically. Handwashing was described as inconvenient and less of a priority for their time over other activities that need their attention, particularly for mothers.

The most commonly reported enablers in domestic settings related to *Reflective* (‘perceived health risk’ (24/54; 44%)), and *Automatic Motivation* (‘internal motivation/habit’ (19/54, 35%)). Study findings show that when people identified a risk to their health, they were motivated to start, maintain, or increase the frequency with which they practice handwashing. Similarly, perceiving handwashing to be good for health also influenced the behavior (high confidence). Regarding ‘internal motivation/habit’ studies show that an established habit, routine, or perceived ‘need’ to wash hands influence practice, and that these routines, habits, and needs were often described in relation to other preceding activities (like toileting) or feelings (disgust).

Additional information describing how and why other sub-themes are noted to be barriers and enablers in the domestic setting is provided in S9- Thematic Analysis Table.

### 3.6. COM-B-categorized barriers and enablers of hand hygiene practices in institutional settings

In institutional settings, ‘soap availability’ (*Physical Opportunity;* 7/21, 33%), ‘time prioritization’ (*Reflective Motivation;* 7/15, 33%), and ‘community infrastructure’ (*Physical Opportunity;* 6/21, 29%) were the most frequently reported barriers. As in domestic settings, studies found that the lack of soap availability in institutional settings hindered handwashing practice (high confidence). ‘Time prioritization’, specifically time pressure, being busy, and having competing priorities or interests—for example kids preferring to use time to play over waiting in a queue while at school—also limited handwashing (high confidence). Finally, both a lack of infrastructure or having limited access to infrastructure in institutional settings were described to hinder behavior (high confidence).

The most commonly reported enablers in institutional settings were ‘internal motivation/habit’ (*Automatic Motivation;* 6/21, 29%), ‘action knowledge’ (*Psychological Capability;* 6/21, 29%), and perceived health risk’ (*Reflective Motivation;* 5/21, 24%). The presence of a habit was described as driving handwashing habits in institutional settings, though studies were limited in both geographic (half in US) and setting scope (mostly schools) (moderate confidence). ‘Action knowledge’, specifically practical knowledge about when and how to wash hands was reported in studies to influence handwashing, though studies were mostly in education settings (schools, universities) and had limited geographic scope (moderate confidence). Finally, regarding ‘perceived health risk’, studies reported handwashing behavior to be influenced both by whether individuals perceived their health to be at risk or the health of others (e.g., customers in a workplace setting) if they do not wash their hands. The theme is predominantly informed by studies in Africa and in education settings (moderate confidence).

Additional information describing how and why other sub-themes are noted to be barriers and enablers in institutional settings is provided in S9- Thematic Analysis Table.

### 3.6. COM-B-categorized barriers and enablers of hand hygiene practices in public settings

In public settings, the most frequently reported barriers were ‘soap availability’ (*Physical Opportunity;* 6/15, 40%), ‘time prioritization’ (*Reflective Motivation;* 5/15, 33%), ‘internal motivation/habit’ (*Automatic Motivation;* 4/15, 27%). As with both domestic and institutional settings, lack of access to soap was found to hinder handwashing practice though most of the small number of studies (5 of 6) were in Africa, of which half were focused on IDP camps, limiting information from other geographies and public settings (moderate confidence). Regarding ‘time prioritization’ being busy or having competing priorities or interests were noted to hinder handwashing at all or as noted in guidelines. Specifically, one study noted that participants were reluctant to follow guidelines because they felt the process to be prolonged if they were to rub and use soap as directed (moderate confidence). Finally, in terms of ‘internal motivation’ participants’ cited their handwashing to be hindered by a lack of habit, routine, or ability to remember to wash their hands, as well as the presence of other routines that do not include handwashing. A participant from one study at a harm reduction center in France summarized how age can also be a factor in whether or not habits can evolve: ‘it’s good for the new generations who are going to adopt this practice. It’s more complicated for older generations” (Mezaache 2021).

The most common enablers in public settings were ‘feeling of cleanliness’ (*Automatic Motivation;* 4/15, 27%), ‘like/dislike of handwashing product’ (*Automatic Motivation;* 3/15, 14%) and ‘perceived health risk’ (*Reflective Motivation;* 3/15, 14%). While these were the most common enablers, confidence in evidence for the themes was either low or very low; the limited number of studies contributed to lack of coherence, adequacy, and relevance. Regarding ‘Feeling of cleanliness’, included studies reported handwashing being motivated because the practice made participants feel clean and refreshed, and for ‘like/dislike of handwashing product’, the feeling of the product (e.g., not greasy), the smell, and how it made hands feel (e.g., feel soft, smooth), inspired use. Finally, as in other settings, illness prevention was reported to drive handwashing.

Additional information describing how and why other sub-themes are noted to be barriers and enablers in public settings is provided in S9- Thematic Analysis Table.

## 4. DISCUSSION

Our review of hand hygiene barriers in community settings relied on qualitative data to ensure understanding of the factors that influence behavior from individual perspectives. We found handwashing with soap and water to be the most dominant hand hygiene behavior assessed. Most of the research took place in Africa and Southeast Asia and in domestic settings and in rural areas, with limited research in workplaces and public spaces. No studies were identified that focused on persons with disabilities. Notably, across all settings, a lack of soap and water (‘Physical Opportunity’) was a dominant barrier, underscoring how vital these resources are to practicing hand hygiene. Even where water and soap were available, constrained access to these resources, including high perceived costs of soap or distance to water sources, was reported to impact regular practice of hand hygiene. ‘Perceived health risk’ and ‘time prioritization’ (‘Reflective motivation’) and the presence (or absence) of a habit (‘Automatic Motivation’) are other highly reported barriers/enablers that should be considered, based on context, once ensuring that water, soap, or other enhancements to the physical environment are in place. Below, we discuss these key findings related to the dominant barriers and enablers identified across settings.

### Access to soap and water is essential but alone likely insufficient to motivate or sustain hand hygiene

Consistent with other reviews ^18,19^, our findings confirm that a lack of an enabling physical environment— that is, one that provides the resources necessary—for hand hygiene like water and soap--can hinder hand hygiene behaviors. Specifically, a 2023 systematic review, which sought to determine the barriers and enablers specific to handwashing with soap in community settings (46 studies), found ‘environmental context and resources’ to be the most common domain described. The authors’ description of this domain aligns with what is referred to in this review as ‘Physical Opportunity’. (Like ‘Physical Opportunity’, the 2023 review authors note that their ‘environmental context and resources’ domain includes a lack of handwashing resources, like handwashing station hardware, water, and soap ^19^). Collectively, these findings underscore the imperative for people to have access to water and soap and/or alcohol-based hand rubs to practice hand hygiene. Even beyond the sphere of hand hygiene, making changes to environments is understood to be crucial to facilitating positive behaviors. Specifically, interventions that change the environmental context not only ‘require less individual effort and have the greatest population impact’ but also help to make the behavior a ‘default choice’ ^45^. Yet, despite the obvious need for access to soap and water (or other hand hygiene materials like alcohol-based hand rubs), another recent review found that interventions designed to improve hand hygiene in community settings are not always providing these necessary resources. Instead, interventions largely focused on providing instructions on how to wash hands (an example of ‘Action knowledge’ under *‘Psychological Capability*’) ^46^. In our review, ‘Psychological Capability’ was not a dominant barrier or enabler, except in school settings where ‘action knowledge’ was noted as an enabler, particularly among school children who may have been learning how to perform the behavior correctly for the first time. Critically, initiatives that aim to improve hand hygiene in community settings—whether in domestic, institutional, or public spaces—first need to enhance the environment with the resources required for handwashing or ensure that they are already in place either before or simultaneous to addressing other barriers or enablers.

### Motivation influences hand hygiene and needs to be considered alongside resource provision

A *‘Reflective Motivation’* theme, ‘time prioritization’, was found to hinder behavior; similar concerns about not having time for handwashing or deeming other activities to be priorities was also noted in other reviews ^18,19^. Specifically, when individuals had competing priorities, interests, or needs, or felt that handwashing—or accessing the materials necessary for handwashing—was too time consuming, they were deterred from practicing the behavior. Following from what was noted previously, individuals may feel that handwashing is not ‘as good’ or ‘as important’ as other behaviors or practices that require their limited time and attention. As such, in addition to elevating what is ‘good’ about handwashing, initiatives can consider how to improve self-efficacy so that handwashing is not believed to be a challenging, time consuming task in competition with other behaviors. Initiatives should ensure that resources are not just available in the environment but easily accessible and conveniently located to minimize or eliminate the time burden and individuals’ associated beliefs that handwashing is inconvenient and they are not able to perform it. Lessons can be learned from research in health care settings, which has found that increasing accessibility and visibility of hand hygiene stations improved handwashing behaviors ^47,48^. Additionally, interventions could ‘bundle’ hand hygiene *with* relevant priority behaviors, so it is considered as a fundamental component of key tasks they would not compromise. For example, interventions have effectively bundled handwashing with child feeding into a ‘mealtime’ package, so individuals do not see handwashing as separate from feeding their children, but as a part of the feeding routine ^49^.

### Building habits is key to sustaining hand hygiene behaviors, provided hygiene resources are available

When people perceived there to be a health risk, they were driven to start handwashing, maintain their practice, or increase the frequency of existing handwashing behaviors. This other *‘Reflective Motivation’* finding may help explain variation in handwashing behavior when health risks were perceived to be more or less acute. For example, a study involving over 6000 adults across 14 countries on five continents found there to be higher levels of handwashing adherence when COVID-19 cases were on the rise in the previous two weeks. Conversely, handwashing fell when COVID-19 cases were not noticeably on the rise, or ‘salient’, in the previous two weeks, but were still gradually accumulating ^50^. The authors observed that this ‘falling and peaking’ pattern of handwashing behavior continued depending on the trajectory of the virus, and that people did not experience a ‘pandemic fatigue’ that eventually led them to stop washing hands and never start up again. During the ‘fall’ part of the cycle, individuals may have lost the belief or perception that the health risk was acute, leading them to believe that handwashing was no longer an urgent need. Important, though, is that the authors found that the cycle would ‘peak’ again when the perception of risk became more salient during which time people would resume more adherent handwashing behaviors. As it would not be advisable to try to maintain people’s concern about the intensity of acute health risks simply to drive handwashing behavior, interventions can work to enshrine the belief or judgement that adherence to handwashing is a ‘good’ behavior even when risks are less acute.

The theme ‘internal motivation/habit’ (Automatic motivation) was the only sub-theme identified as a dominant barrier and enabler both overall and in domestic and institutional settings. Across various studies, individuals reported how handwashing was easy because they already had a habit or that it was not something they did because they lacked a habit. Different approaches are needed to address ‘Automatic Motivation’ depending on whether habits need to be created or maintained. Establishing new routines and regularly practicing or repeating these routines until they are automatic are seen as critical for habit formation ^51^, as is adding cues to the environment to trigger the behavior or ‘piggy-backing’ on other established routines ^52^. Critically, effort should be made to maintain environments that are supportive of existing habits; when environments that previously supported habits change, habits might be broken ^52^. Specifically, if hand hygiene materials are consistently available to encourage handwashing or sanitizing but then are removed, people may not maintain the habit by seeking out or providing the materials for themselves. In fact, changing the environment, which can disrupt an individual’s routine, is seen as a strategy for breaking bad habits ^52^. Therefore, an unintentional outcome of no longer providing hand sanitizers in public spaces like schools, transport hubs, or shops—as was typical during the height of the COVID-19 pandemic—is that people will no longer practice hand hygiene when entering through a door to a new space. Even worse, the absence of a hand hygiene station may act as its own cue to signal that the behavior is no longer necessary.

Findings underscore the importance of not only establishing habits for hand hygiene but ensuring that habits can be maintained by ensuring that necessary resources are available to enable it (e.g., creating ‘Physical Opportunity’). If resources are removed or are not replenished when needed, behaviors that were well established may be easily forgotten. Further, many studies discussed how people had habits that did not include hand hygiene, or their habit was to not practice hand hygiene. As such, in many cases interventions and programs may need to not only aim to create habits, but they also may need to break existing habits that do not include hand hygiene.

### Hand hygiene interventions must be inclusive, addressing the barriers and enablers for all

In our review, no studies focused on barriers and enablers of hand hygiene practice for people with disabilities. People with disabilities are a key vulnerable group that face specific barriers hindering their hand hygiene practice, with barriers varying by the type of disability ^53^. A study during COVID-19 in five different countries (Kenya, Indonesia, Zambia, Sierra Leone and Bangladesh) shows that people with disabilities faced significant barriers to accessing and using handwashing stations in both household and public settings ^54^. Understanding the varied barriers and enablers for people with disabilities is necessary to ensure that interventions are adequately designed to improve hand hygiene for all.

Behavioral theory was used in studies investigating the drivers and barriers of hand hygiene, though by only a third of studies. As this study aimed only to see how common theory is in the study of hand hygiene in community settings, further research could be undertaken to understand how theory was used (e.g., in design of tools, during analysis, etc.).

### Strengths and Limitations

This review was part of an integrated protocol for multiple related reviews, which included an exhaustive search strategy encompassing multiple databases and grey literature sources, and a two-phased approach to identify relevant literature of hand hygiene in community settings. To understand barriers and enablers, we deliberately sought qualitative research. Qualitative approaches encourage those engaged to voice their own insights and ideas in an open-ended way. Therefore, we expected that the studies included may have captured a broader array of insights regarding hand hygiene behavior than what would have been captured by quantitative studies, which would use researcher-developed surveys to assess the frequency or agreement of barriers and enablers selected a priori. In reviewing and assessing the barriers and enablers described in the studies, we applied the widely-used COM-B framework. While well-regarded in behavior change research, the COM-B model was only used by 5% of the studies included and our categorization of behavioral barriers and enablers may be different than what the primary study authors might have done. Still, the use of COM-B enabled us to compare findings and identify trends across diverse studies. Further, while we did investigate the use of theory among included studies, this study aimed only to see how common it has been to use theory in research on hand hygiene in community settings. It was beyond the scope of this to understand *how* theory was used (e.g., in design of tools, during analysis, etc.) and if done so effectively; further research could be undertaken to explore theory use in more depth. Finally, while this review did not focus solely on handwashing with water and soap, only a limited number of studies (6) explicitly investigated barriers and enablers to hand hygiene with other materials (e.g., water only, soap alternatives, and alcohol-based rub). Because of this limited sample, we are not able to discern if barriers and enablers are different based on hand hygiene methods and encourage further research, particularly into alcohol-based hand rubs which are widely used and were ubiquitous during the height of the COVID-19 pandemic.

## 5. CONCLUSION

Our finding that a lack of access to soap and water was a dominant barrier to hand hygiene but that access to these resources was not a major enabler suggests that soap and water are necessary but may not be sufficient to drive hand hygiene behavior. As such, programs that seek to enable hand hygiene behaviors should first ensure that physical environments are equipped with the resources necessary for handwashing and then look to other behavioral enablers and barriers, with attention to the specific setting and the local context.

## Authors’ contributions

OC, JEM, and BG conceived the review and designed the specific research questions. BAC and MW are the guarantors of the review. BAC, JSS, and LAO conceived and designed the specific analysis strategy and data extraction process described herein. HKR led the literature search process. EL, KF and DMS conducted screenings. LAO, EL, KF and DMS extracted data with BAC and JSS support. LAO developed the data extraction tool informed by and with support from SKP. LAO and JSS led data visualization. DMS assisted with data visuals. BAC and LAO led the analysis. BAC led manuscript writing and oversaw revisions. JSS drafted portions of the manuscript. All authors contributed to the interpretation of the results, and read and approved the final version of the manuscript.

## Acknowledgements

The authors would like to extend their gratitude to Dr. Susan Michie for reviewing and providing edits to the codebook for classifying enablers and barriers according to the COM-B model. The authors thank the team of screeners who contributed substantially to the review process as part of phase 1 and phase 2 screening (Kainalu Bailey, Nick An, Josef Zhao, Filmon Gebremichael, Jordan Honeycutt, Norah McKinley, Erika Canda, Michael Horner-Ibler, Liya Getachew).

