## Supplementary material for "Behavioural factors influencing hand hygiene practices across domestic, institutional, and public community settings: A systematic review": S2

**S2**- Prisma Flow Diagram

1 Hubert Department of Global Health, Rollins School of Public Health, Emory University, Atlanta, GA, USA; (BAC); (DMS); (SKP)
2 Gangarosa Department of Environmental Health, Rollins School of Public Health, Emory University, Atlanta, GA, USA; (MCF); (MW) (JSS); (LAO); Kennedy Files (KF); Erin LaFon (EL)
3 Woodruff Health Sciences Center Library, Emory University, Atlanta, GA, USA; (HR)
4 Department of Disease Control, London School of Hygiene and Tropical Medicine, London, UK; (OC)
5 Water, Sanitation, Hygiene and Health Unit, World Health Organization, Geneva, Switzerland; (JEM)
Emory University, Rollins School of Public Health, 1518 Clifton Rd, Atlanta, GA 30322

*Contributed equally.

**S2 -** PRISMA flow chart.

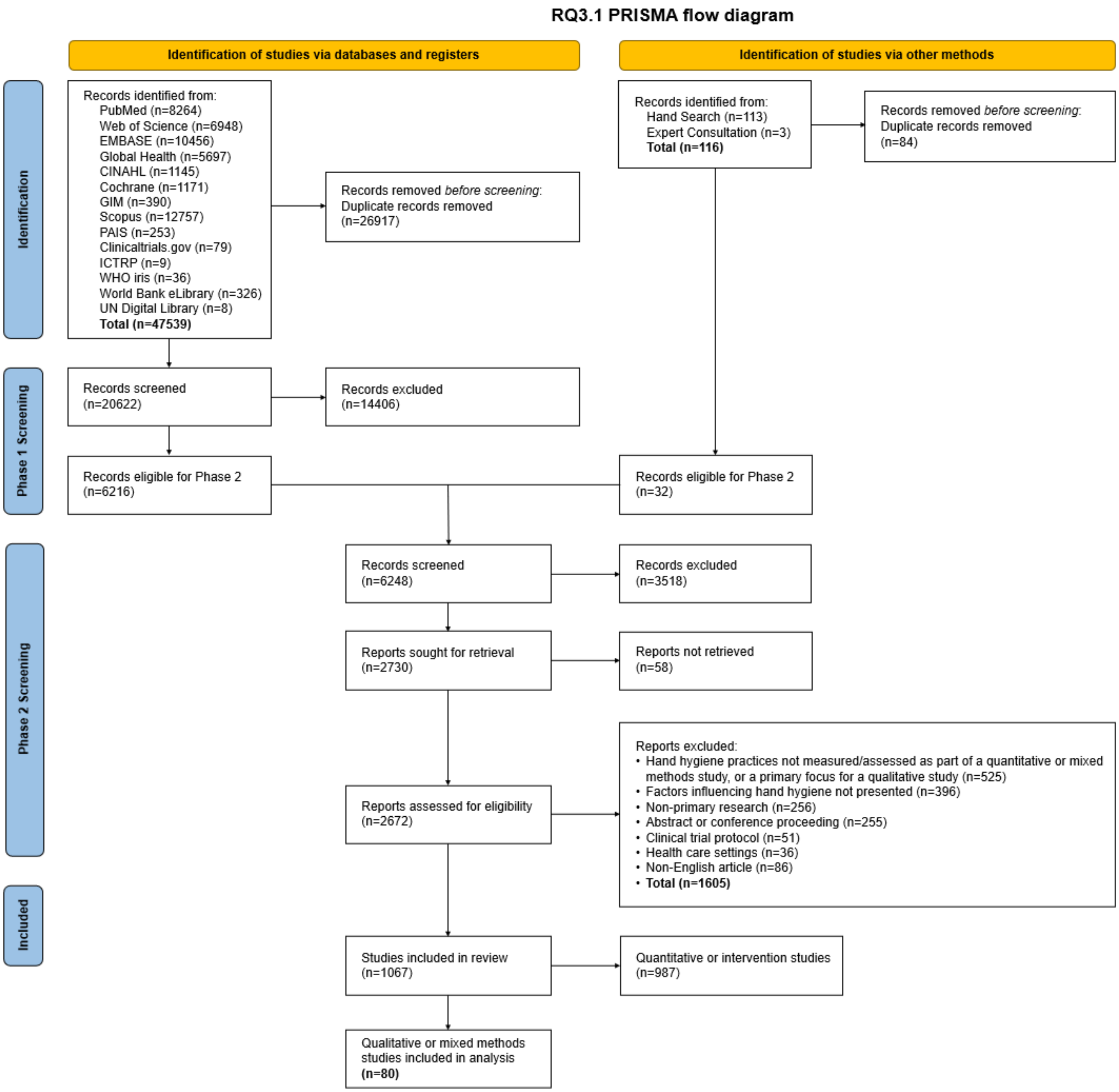
