## Supplementary material for "Behavioural factors influencing hand hygiene practices across domestic, institutional, and public community settings: A systematic review": S3

**S3-** Covidence Extraction Questionnaire

1 Hubert Department of Global Health, Rollins School of Public Health, Emory University, Atlanta, GA, USA; (BAC); (DMS); (SKP)
2 Gangarosa Department of Environmental Health, Rollins School of Public Health, Emory University, Atlanta, GA, USA; (MCF); (MW) (JSS); (LAO); Kennedy Files (KF); Erin LaFon (EL)
3 Woodruff Health Sciences Center Library, Emory University, Atlanta, GA, USA; (HR)
4 Department of Disease Control, London School of Hygiene and Tropical Medicine, London, UK; (OC)
5 Water, Sanitation, Hygiene and Health Unit, World Health Organization, Geneva, Switzerland; (JEM)
Emory University, Rollins School of Public Health, 1518 Clifton Rd, Atlanta, GA 30322

*Contributed equally.

### Extraction fields for RQ3.1: Barriers and Enablers to Hand Hygiene in Community Settings

| | |
|---|---|
| | Item extracted in all RQs |
| | Item specific to Barriers and enablers (3.1) |

| # | Field | Details | Entry |
|---|---|---|---|
| **1. Information about the study** | | | |
| 1.1 | Study design | | *Select one*<br>(1) Qualitative study<br>(2) Mixed methods<br>(3) Non-comparative<br>(4) Experimental Before-and-after study<br>(5) Observational Before-and-after study<br>(6) Experimental Times series<br>(7) Observational Times series<br>(8) Non-randomized trial<br>(9) Non-randomized cross-over design<br>(10) Randomized controlled trial<br>(11) Randomized cross-over study<br>(12) Analytical cross-sectional study<br>(13) Case-control study<br>(14) Cohort study<br>(777) Other – specify |
| 1.2 | Registered trial | Does the study report that it is linked to a registered trial? (e.g., clinicaltrials.gov, ICTRP) | *Select one*<br>(1) Yes<br>(0) No |
| 1.3 | Trial # | If linked to a registered trial, paste trial registration number | Text<br>(999) Not applicable |
| **2. Eligibility** | | | |
| 2.1 | Confirm participants/sample | Does the study include general populations in community settings? | *Select one*<br>(1) Yes<br>(0) No |

| | | | |
|---|---|---|---|
| | | DO NOT PROCEED IF "NO" | |
| 2.2 | Confirm phenomena of interest | Is the phenomena of interest "behavioral barriers and enablers for practicing hand hygiene?"<br><br>DO NOT PROCEED IF "NO" | *Select one*<br>(1) Yes<br>(0) No |
| 2.3 | Confirm study design | Is the design of the study NOT a quantitative non-comparative study?<br><br>DO NOT PROCEED IF "NO" | *Select one*<br>(1) Yes<br>(0) No |
| 2.4 | Confirm evaluation | Does the study include the evaluation of hand hygiene practice (i.e., any action of hand cleansing for the purpose of removing or deactivating pathogens from hands)?<br><br>DO NOT PROCEED IF "NO". | *Select one*<br>(1) Yes<br>(0) No |
| 2.5 | Confirm research type | What research type does the study use?<br><br>DO NOT PROCEED IF NONE ARE CHECKED | *Select one*<br>(1) Qualitative<br>(2) Quantitative<br>(3) Mixed methods |
| **3. Setting** | | | |
| 3.1 | Country | Which country is represented in the study? (List all countries separated by a comma, if study is from multiple sites) | Text<br>999 = Not applicable |
| 3.2 | Region | Which region is represented in the study? | *Check multiple*<br>(1) Africa<br>(2) Asia<br>(3) Europe<br>(4) Latin America/Caribbean<br>(5) Middle East<br>(6) North America |

| | | | |
|---|---|---|---|
| | | | (8) Oceania<br>(10) Unspecified<br>(999) Not applicable |
| 3.3 | Urban/Rural | Does the setting of the population fall under any of these specific categories? Select all that apply | *Check multiple*<br>(1) Urban<br>(2) Rural<br>(3) Peri-urban<br>(777) Other - specify<br>(888) Not reported |
| 3.4 | Community setting | Does the setting of the population fall under any of these specific categories? Select all that apply | *Check multiple*<br>(1) Domestic - Households<br>(2) Public - Markets<br>(3) Public - Public transportation hubs<br>(4) Public - Parks, squares, or other public outdoor spaces,<br>(5) Institutions - Workplace<br>(6) Institutions - Schools<br>(7) Institutions - Universities<br>(8) Institutions - Places of worship<br>(9) Institutions - Prisons and places of detention<br>(10) Internally displaced people camps<br>(777) Other - specify<br>(888) Not reported |
| 3.5 | Risk scenarios | Did the study evaluate hand hygiene in a risk scenario? | *Select one*<br>(0) No<br>(1) Yes<br>(999) Unclear |
| 3.6 | Risk scenario type | If yes, what was the risk scenario? | *Select one*<br>(1) COVID-19<br>(2) Flu<br>(3) Earthquake<br>(4) Flood<br>(5) Typhoon<br>(6) Forced migration<br>(7) Internal displacement<br>(8) Emergency setting<br>(777) Other – specify |

| | | | |
|---|---|---|---|
| | | | (999) Not applicable |
| 3.7 | Risk scenario text | Please copy in the author's text about the risk scenario. | Text<br>(999) Not applicable |
| **4. Methods** | | | |
| 4.1 | Aim of study | Copy and paste the aim/objective/purpose/goal as stated in the study | Text<br>(888) Not reported |
| 4.2 | Primary study outcome | What was the primary outcome for this study?<br>Select all that apply | *Check multiple*<br>(1) Hand hygiene<br>(2) Diarrheal diseases<br>(3) Respiratory infections<br>(4) Influenza<br>(5) Other Infectious diseases<br>(6) Nutrition<br>(7) Mental/social well being<br>(8) Neglected tropical diseases<br>(9) School absenteeism<br>(10) COVID-19<br>(11) Food hygiene<br>(12) Soil-transmitted helminth infection<br>(777) Other – specify<br>(999) Not applicable |
| 4.3 | Start date | What is the study start date? Month, Year | Text<br>(888) Not reported<br>(999) Not applicable |
| 4.4 | End date | What is the study end date? Month, Year | Text<br>(888) Not reported<br>(999) Not applicable |
| **5. Participants** | | | |
| 5.1a | Study participants-Health outcome | What group(s) of people are researchers examining the focal health outcome for in the study?<br>Select all that apply | *Check multiple*<br>(1) General population<br>(2) Adults (Women and Men)<br>(3) Adults (Women only)<br>(4) Adults (Men only)<br>(5) Children (Girls and Boys)<br>(6) Children (Girls only) |

| | | | |
|---|---|---|---|
| | | | (7) Children (Boys only)<br>(8) Mother-child dyads<br>(9) Food workers<br>(10) Non-food occupational workers<br>(777) Other – specify<br>(888) Not reported<br>(999) Not applicable |
| 5.1b | Study participants-Hand washing behavior | What group(s) of people are researchers examining hand washing behavior for in the study?<br>Select all that apply | *Check multiple*<br>(1) General population<br>(2) Adults (Women and Men)<br>(3) Adults (Women only)<br>(4) Adults (Men only)<br>(5) Children (Girls and Boys)<br>(6) Children (Girls only)<br>(7) Children (Boys only)<br>(8) Mother-child dyads<br>(9) Food workers<br>(10) Non-food occupational workers<br>(777) Other – specify<br>(888) Not reported<br>(999) Not applicable |
| 5.2 | Vulnerable populations | Does study concern any of the following vulnerable populations?<br>Select all that apply | *Check multiple*<br>(1) Individuals with specific illness or risk factors<br>(2) Specific ethnic or religious groups<br>(3) Persons experiencing homelessness<br>(4) Persons with disabilities<br>(5) Immigrants and migrants<br>(6) Refugees and displaced persons (7) Elderly<br>(8) Pregnant women<br>(777) Other - specify<br>(888) Not reported |
| 5.3 | Number of participants | What is the total number of participants/sample size? | Table<br>2 x 3 table for Adult/Child vs Female/Male/Total |

| | | | |
|---|---|---|---|
| | | If the study does not stratify participants by sex or gender, only fill in the total row. If does not stratify by adult/child, only fill in adult column. | |
| 5.4 | Number of participants (mixed methods) | For mixed methods studies, how many participants are involved with each method? | Table<br>3x2 table for Qual/Quant/Total |
| **6. Intervention** | | | |
| 6.1 | Theory | Did the study report using a behavior change theory? Select all that apply (model, framework) | *Select multiple*<br>(1) IBM-WASH<br>(2) RANAS<br>(3) Behavior Centered Design/Evo-Eco Model<br>(4) COM-B<br>(5) Theory of Planned Behavior<br>(6) Health Belief Model<br>(7) Social Ecological Model<br>(8) Theoretical Domains Framework<br>(777) Other – specify<br>(999) No theory reported |
| 6.2 | Theory mention | Where did the authors discuss the theory that they used? | *Select one*<br>(1) Protocol<br>(2) Cited formative research<br>(3) Primary research paper<br>(777) Other – specify<br>(999) No theory reported |
| **7. Outcomes (RQ3.1)** | | | |
| 7.1 | Physical Capability | Were physical capability barriers or enablers presented in the study?<br><br>Definition of physical capability: physical strength, skill, stamina | (1) Yes<br>(0) No |

| | | | |
|---|---|---|---|
| 7.2 | Psychological Capability | Were psychological capability barriers or enablers presented in the study?<br><br>Definition of psychological capability: knowledge/psychological strength, skills or stamina | (1) Yes<br>(0) No |
| 7.3 | Social opportunity | Were social opportunity barriers or enablers presented in the study?<br><br>Definition of social opportunity: opportunities as a result of social factors, such as cultural norms and social cues | (1) Yes<br>(0) No |
| 7.4 | Physical opportunity | Were physical opportunity barriers or enablers presented in the study?<br><br>Definition of physical opportunity: opportunities provided by the environment, such as time, location and resources | (1) Yes<br>(0) No |
| 7.5 | Automatic motivation | Were automatic motivation barriers or enablers presented in the study?<br><br>Definition of automatic motivation: automatic processes, such as our desires, impulses and inhibitions | (1) Yes<br>(0) No |
| 7.6 | Reflective motivation | Were reflective motivation barriers or enablers presented in the study? | (1) Yes<br>(0) No |

| | | |
|---|---|---|
| | | Definition of reflective motivation: reflective processes, such as making plans and evaluating things that have already happened |

#### Bias assessment

| **Bias Assessment- MMAT** | | | | |
|---|---|---|---|---|
| All articles | | | | |
| **#** | **Field** | **Details** | **Entry** | **Source** |
| S1 | Screening question 1 (for all types) | Are there clear research questions? | (0) No<br>(1) Yes<br>(999) Can't tell | MMAT User Guide |
| S2 | Screening question 2 (for all types) | Do the collected data allow to address the research questions? | (0) No<br>(1) Yes<br>(999) Can't tell | MMAT User Guide |
| Qualitative | | | | |
| 1.1 | Is the qualitative approach appropriate to answer the research question? | | (0) No<br>(1) Yes<br>(999) Can't tell | MMAT User Guide |
| 1.2 | Are the qualitative data collection methods adequate to address the research question? | | (0) No<br>(1) Yes<br>(999) Can't tell | MMAT User Guide |
| 1.3 | Are the findings adequately derived from the data? | | (0) No<br>(1) Yes<br>(999) Can't tell | MMAT User Guide |
| 1.4 | Is the interpretation of results sufficiently substantiated by data? | | (0) No<br>(1) Yes<br>(999) Can't tell | MMAT User Guide |
| 1.5 | Is there coherence between qualitative data sources, collection, analysis and interpretation? | | (0) No<br>(1) Yes<br>(999) Can't tell | MMAT User Guide |
| Quantitative randomized controlled trials | | | | |
| 2.1 | Is randomization appropriately performed? | | (0) No<br>(1) Yes | MMAT User Guide |

| | | | |
|---|---|---|---|
| | | (999) Can't tell | |
| 2.2 | Are the groups comparable at baseline? | (0) No<br>(1) Yes<br>(999) Can't tell | [MMAT User Guide] |
| 2.3 | Are there complete outcome data? | (0) No<br>(1) Yes<br>(999) Can't tell | [MMAT User Guide] |
| 2.4 | Are outcome assessors blinded to the intervention provided? | (0) No<br>(1) Yes<br>(999) Can't tell | [MMAT User Guide] |
| 2.5 | Did the participants adhere to the assigned intervention? | (0) No<br>(1) Yes<br>(999) Can't tell | [MMAT User Guide] |
| Quantitative non-randomized | | | |
| 3.1 | Are the participants representative of the target population? | (0) No<br>(1) Yes<br>(999) Can't tell | [MMAT User Guide] |
| 3.2 | Are measurements appropriate regarding both the outcome and intervention (or exposure)? | (0) No<br>(1) Yes<br>(999) Can't tell | [MMAT User Guide] |
| 3.3 | Are there complete outcome data? | (0) No<br>(1) Yes<br>(999) Can't tell | [MMAT User Guide] |
| 3.4 | Are the confounders accounted for in the design and analysis? | (0) No<br>(1) Yes<br>(999) Can't tell | [MMAT User Guide] |
| 3.5 | During the study period, is the intervention administered (or exposure occurred) as intended? | (0) No<br>(1) Yes<br>(999) Can't tell | [MMAT User Guide] |
| Quantitative descriptive | | | |
| 4.1 | Is the sampling strategy relevant to address the research question? | (0) No<br>(1) Yes<br>(999) Can't tell | [MMAT User Guide] |
| 4.2 | Is the sample representative of the target population? | (0) No<br>(1) Yes<br>(999) Can't tell | [MMAT User Guide] |

| | | | |
|---|---|---|---|
| 4.3 | Are the measurements appropriate? | (0) No<br>(1) Yes<br>(999) Can't tell | MMAT User Guide |
| 4.4 | Is the risk of nonresponse bias low? | (0) No<br>(1) Yes<br>(999) Can't tell | MMAT User Guide |
| 4.5 | Is the statistical analysis appropriate to answer the research question? | (0) No<br>(1) Yes<br>(999) Can't tell | MMAT User Guide |
| Mixed methods | | | |
| 5.1 | Is there an adequate rationale for using a mixed methods design to address the research question? | (0) No<br>(1) Yes<br>(999) Can't tell | MMAT User Guide |
| 5.2 | Are the different components of the study effectively integrated to answer the research question? | (0) No<br>(1) Yes<br>(999) Can't tell | MMAT User Guide |
| 5.3 | Are the outputs of the integration of qualitative and quantitative components adequately interpreted? | (0) No<br>(1) Yes<br>(999) Can't tell | MMAT User Guide |
| 5.4 | Are divergences and inconsistencies between quantitative and qualitative results adequately addressed? | (0) No<br>(1) Yes<br>(999) Can't tell | MMAT User Guide |
| 5.5 | Do the different components of the study adhere to the quality criteria of each tradition of the methods involved? | (0) No<br>(1) Yes<br>(999) Can't tell | MMAT User Guide |
