## Supplementary material for "Behavioural factors influencing hand hygiene practices across domestic, institutional, and public community settings: A systematic review": S5

**S5**- COM-B Codebook

1 Hubert Department of Global Health, Rollins School of Public Health, Emory University, Atlanta, GA, USA; (BAC); (DMS); (SKP)
2 Gangarosa Department of Environmental Health, Rollins School of Public Health, Emory University, Atlanta, GA, USA; (MCF); (MW) (JSS); (LAO); Kennedy Files (KF); Erin LaFon (EL)
3 Woodruff Health Sciences Center Library, Emory University, Atlanta, GA, USA; (HR)
4 Department of Disease Control, London School of Hygiene and Tropical Medicine, London, UK; (OC)
5 Water, Sanitation, Hygiene and Health Unit, World Health Organization, Geneva, Switzerland; (JEM)
Emory University, Rollins School of Public Health, 1518 Clifton Rd, Atlanta, GA 30322

*Contributed equally.

### COM-B Codebook

| MAXQDA Code | # | (choose the -00 level when unable to fit into a sub-category) | |
|---|---|---|---|
| *c-phys* | **100** | **Physical Capability** | **A capability that involves a person's physique and musculoskeletal functioning (e.g. brain and extremity)** |
| *c-phys: ability* | | physical ability | Physical limitations that the participant may experience that affect their ability to perform the hand hygiene behavior. (For example, an elderly person may find it difficult to stand for a long time to wash hands, and thus prefers to use hand sanitizer.) |
| | **101** | Ease/difficulty of washing hands | The perceived level of ease or difficulty individuals experience when performing handwashing actions due to individual physical abilities. |
| | **102** | Ease/difficulty of collecting water | The level of physical ease or difficulty individuals face in collecting water for handwashing purposes, which can affect the frequency and quality of handwashing practices. |
| *c-psyc* | **200** | **Psychological Capability** | **A capability that involves a person's mental functioning (e.g. understanding and memory)** |
| *c-psyc: knwldg* | | action knowledge | Participant has or does not have the knowledge necessary to perform the hand hygiene behavior. (For example, knowledge around WHEN it is important to wash one's hands.) |
| *o-phys* | **300** | **Physical Opportunity** | **An opportunity that involves inanimate parts of the environmental system and time (e.g. financial and material resources)** |
| *o-phys: space* | | space/land | Refers to issues around having enough space or land availability in order to practice hand hygiene. (For example, having a suitable area for handwashing facilities.) |
| | **301** | Enough space for a station | The availability or lack of sufficient space to accommodate handwashing stations |
| | **302** | Ownership of land/house | The legal possession or ownership of land or housing, which can influence the feasibility and placement of handwashing facilities |
| *o-phys: envir* | | environment | Refers to the physical environment in which people are living. This may include references to water sources, seasons, etc. ("I cannot wash my hands because the water is too cold during winter.") |
| | **310** | Water quality | The perceived cleanliness and safety of the water used for handwashing |

| | | | |
|---|---|---|---|
| | **311** | Water supply quantity | The quantity of water present at the source used for handwashing water. For example, low levels of water in a well affecting the ability of a household to gather water to wash hands. |
| | **312** | Cleanliness of handwashing environment | The overall cleanliness and hygiene of the area designated for handwashing impacting ability and desire to wash hands in that location. |
| | **313** | Lighting of handwashing area | The presence or absence of adequate lighting in the handwashing area. Can impact visibility of handwashing stations and materials, perception of safety, etc. that affect handwashing behavior |
| | **314** | Temperature of water | The temperature of the water used for handwashing, with considerations for comfort and perceived effectiveness in removing contaminants. |
| | **315** | State of hand hygiene equipment | The functionality or state of repair of hand hygiene equipment which can affect the ability of a person to practice hand hygiene behavior |
| *o-phys: sustain* | | sustainability | Whether the materials, goods, or services for hand hygiene are long-lasting. This may refer to handwashing facilities that are easily damaged during harsh weather conditions. |
| | **330** | Damage of resources | The extent of damage or degradation of resources, such as handwashing facilities, water sources, or soap dispensers, which may impact hand hygiene practices. |
| *o-phys: access* | | accessibility | Whether materials, goods, and services for hand hygiene are close by and easy to get. May be impacted by weather/season, etc. |
| | **340** | Water distance | The physical distance individuals need to cover to access water for handwashing, which can influence the convenience and frequency of hand hygiene. |
| | **341** | Hand hygiene station location | The proximity or distance of handwashing facilities from individuals, affecting the likelihood of use. |
| | **342** | Hand hygiene station design | The design and layout of handwashing facilities, including factors such as accessibility, usability, and hygiene considerations. |
| *o-phys: avail* | | availability | Whether the materials, goods, or services for hand hygiene are available. This may refer to the stock of products at the nearby store (For example, soap) |
| | **360** | Water availability | The quantity, consistency, or reliability of water for handwashing purposes |

| | | | |
|---|---|---|---|
| | **361** | Soap availability | The quantity, consistency, or reliability of soap or cleansing agents for handwashing |
| | **362** | Theft of resources | The occurrence or frequency of theft or unauthorized removal of handwashing-related resources, such as soap or water, which may affect hand hygiene practices. |
| | **363** | Community infrastructure | The presence and condition of community-level infrastructure, including facilities that support hand hygiene, such as communal water sources or public restrooms. |
| | **364** | Household infrastructure | The availability and condition of infrastructure within individual households that supports hand hygiene, such as private handwashing facilities or plumbing. |
| | **365** | Resource availability for purchase | The availability of handwashing-related resources for purchase locally |
| | **366** | Resource provision by external organisation/authority | Support or provision of handwashing-related resources by external organizations or authorities, contributing to the availability and accessibility of such resources. |
| | **367** | Latrine ownership | The ownership and accessibility of latrine facilities contributing to hand hygiene behavior. For example, a household that owns a latrine has the authority to install a handwashing station. |
| *o-phys: afford* | | affordability | Affordability of materials, goods, and services for hand hygiene. Includes references to when things are also not affordable or free of cost. |
| | **380** | Cost of water | The financial burden or cost associated with obtaining water for handwashing, which may impact the affordability and accessibility of hand hygiene practices. |
| | **381** | Cost of soap | The financial cost of purchasing soap or cleansing agents for handwashing, which can influence the affordability and accessibility of hand hygiene practicies. |
| | **382** | Handwash station maintenance cost | The financial expenses associated with maintaining handwashing stations, including repairs, cleaning supplies, and other ongoing costs. |
| *o-soc* | **400** | **Social Opportunity** | **An opportunity that involves other people and organizations (e.g. social and cultural norms)** |
| *o-soc: norm* | | social norm | Participant believes they should practice hand hygiene because others in their reference network conform to the behavior AND they believe that others think they SHOULD perform hand hygiene (normative expectations). Also applies in reverse for not handwashing. |

| | | | |
|---|---|---|---|
| | **420** | Parents model a behavior norm | The behavior of parents or caregivers serving as role models for proper hand hygiene practices, influencing the adoption of similar behaviors by children. |
| | **421** | Social pressure | The influence exerted by social networks or communities to conform to established norms and expectations regarding hand hygiene practices. |
| | **422** | Role modeling | The practice of individuals or community leaders serving as role models for proper hand hygiene, influencing others to follow suit. |
| *o-soc: conseq* | | social consequences | Participant describes a social response (positive or negative) to practicing or not practicing hand hygiene. |
| | **440** | Visits by authorities | Official visits or inspections by authorities, which may influence compliance with hand hygiene practices within a community. |
| | **441** | Exclusion/inclusion by peers | Belief that an individual's hand hygiene behaviors will result in a changed level of inclusion or exclusion in peer groups |
| | **442** | Social stigmatization/status | Belief that an individual's hand hygiene behaviors will result in a changed social status (increased status or decreased status/stigmatization) |
| *o-soc: support* | | social support | The participant describes a provision of assistance or comfort from others. This can be in the form of Information, emotional, tangible/instrumental, network, or esteem support. |
| | **460** | Informational support | Provision of advice, guidance, or information related to hand hygiene practices, assisting individuals in making decisions about hand hygiene behaviors |
| | **461** | Emotional support | Provision of empathy, understanding, and comfort related to hand hygiene, offering individuals emotional reassurance and encouragement. |
| | **462** | Tangible/instrumental support | Provision of practical assistance or resources to facilitate hand hygiene practices, such as providing soap or assisting with handwashing station maintenance. |
| | **463** | Network support | Presence or absence of social connections or community networks that influence hand hygiene practices |
| | **464** | Esteem support | Enhancement of individuals' self-esteem and confidence in their ability to practice behaviors, through recognition, praise, or positive regard from others in their social environment. |
| *o-soc: culture* | | "our culture" | Refers to perceived descriptive norms - behaviors that individuals prefer to conform to on the condition that they |

| | | | |
|---|---|---|---|
| | | | believe most people in their reference network conform to it. In other words, this code is all about people performing hand hygiene because OTHERS around them perform hand hygiene as well. This code can be applied when it is clear that the participant is performing hand hygiene because others in their community perform hand hygiene and it is "our culture" but not when the participant actually describes the EXPECTATION that others believe they SHOULD perform hand hygiene otherwise there will be social consequences. |
| | **480** | Cultural practices and beliefs | The influence of cultural practices, traditions, and beliefs on hand hygiene behaviors, shaping individuals' attitudes and approaches to cleanliness. |
| *m-ref* | **500** | **Reflective Motivation** | **The motivation that involves conscious thought processes (e.g. evaluations and plans)** |
| *m-ref: decis* | | decision-making | Description of the decisions that a person makes around hand hygiene practices. (For example, a person decides to prioritize handwashing before meals.) |
| | **501** | Willingness to practice handwashing | The individual's readiness and willingness to engage in handwashing practices, reflecting their attitude toward factors related to hand hygiene |
| | **502** | Responsibility ownership in maintaining hand hygiene spaces | The presence or absence of a sense of responsibility or ownership among individuals or communities to ensure the upkeep and maintenance of handwashing materials |
| *m-ref: priority* | | competing priorities | When a person puts more importance on one practice/behavior over another. (For example, a person will not use water for handwashing because they need it for cooking. "I will save my water to cook/drink, not to wash my hands.") —> If time is being prioritized, choose "time" |
| | **551** | Soap prioritization | The level of prioritization of soap or cleansing agents for hand hygiene purposes in resource allocation or budgeting (for example, one may prioritize soap for laundry over handwashing) |
| | **552** | Water prioritization | The level of prioritization of water resources for hand hygiene purposes in resource allocation or budgeting (for example, one may prioritize water for cooking over handwashing) |
| | **553** | Time prioritization | The level of prioritization of time for practicing hand hygiene compared to other behaviors |
| *m-ref: risk* | | perceived severity/risk | Participant's perception of how vulnerable they are to health issues (risk) and/or how serious is the health issue (severity). Relates to the health problem, not the hand hygiene behavior. |

| | | | |
|---|---|---|---|
| | | | ("I don't need to worry about germs because that's only a health concern for children.") |
| | **561** | Disease risk | The perceived risk or seriousness of potential diseases or illnesses resulting from hand hygiene behaviors, which may motivate individuals to engage in specific hand hygiene behaviors |
| *m-ref: diffic* | | perceived difficulty | Participant's belief of how difficult it is to perform hand hygiene (whether it is easy, difficult, or neither easy nor difficult, etc.) |
| | **571** | Ability to maintain cleanliness | The perceived level of ease or difficulty individuals associate with maintaining cleanliness or engaging in handwashing practices, which can influence motivation and adherence to hand hygiene behaviors |
| | **572** | Perceived ease/difficulty of washing hands | The perceived level of ease or difficulty individuals associate with washing their hands, which can influence motivation and adherence to hand hygiene behaviors |
| *m-ref: self-eff* | | self-efficacy | Participant's belief that they can or cannot practice hand hygiene, or have the ability/do not have the ability to do something. |
| | **581** | self-efficacy | The belief in one's ability to perform handwashing actions effectively and consistently, reflecting confidence in maintaining proper hand hygiene habits. |
| *m-ref: divine* | | divine influence | Role of religion around why or how hand hygiene behaviors are performed. Can also include participant discussion of 'divine will' - the belief that illness is God's will. Includes the participant's perception of what their religion accepts or rejects and perceptions about the spirit world or magic (e.g., spells, curses). |
| | **591** | God determines sickness | The belief that sickness and health outcomes are determined by a higher power or divine influence, which may affect attitudes and practices related to hand hygiene. |
| *m-auto* | **600** | **Automatic Motivation** | **The motivation that involves habitual, instinctive, drive related, and affective processes (e.g. desires and habits)** |
| *m-auto: attitude* | | attitudes | Participant's attitude towards hand hygiene including whether or not they like or dislike the behavior, feelings, and emotions they associate with hand hygiene ("I enjoy the freshness after washing hands"). This code also includes personal norms - whether or not the participant believes hand hygiene SHOULD be practiced (approval/disapproval of the behavior). |

| | | | |
|---|---|---|---|
| | **601** | Like/dislike the handwash material smell | Individual attitudes or preferences regarding handwashing materials, such as liking or disliking the smell or feel of soap, which may influence motivation to engage in hand hygiene. |
| | **602** | Like/dislike the handwash material feeling | Individuals' preference or aversion to the tactile sensation or texture of handwashing materials, such as soap, influencing their motivation to engage in hand hygiene practices. |
| | **603** | Feeling of cleanliness | The subjective perception or sensation experienced after handwashing, often referring to the immediate sensory experience or emotional responses, which may influence individuals' satisfaction with the hand hygiene process. |
| | **604** | Internal motivation | Internal factors or psychological states such as inertia or lack of motivation that may hinder individuals from consistently practicing hand hygiene despite understanding its importance and benefits. |
