## Supplementary material for "Behavioural factors influencing hand hygiene practices across domestic, institutional, and public community settings: A systematic review": S6

1 Hubert Department of Global Health, Rollins School of Public Health, Emory University, Atlanta, GA, USA; (BAC); (DMS); (SKP)
2 Gangarosa Department of Environmental Health, Rollins School of Public Health, Emory University, Atlanta, GA, USA; (MCF); (MW) (JSS); (LAO); Kennedy Files (KF); Erin LaFon (EL)
3 Woodruff Health Sciences Center Library, Emory University, Atlanta, GA, USA; (HR)
4 Department of Disease Control, London School of Hygiene and Tropical Medicine, London, UK; (OC)
5 Water, Sanitation, Hygiene and Health Unit, World Health Organization, Geneva, Switzerland; (JEM)
Emory University, Rollins School of Public Health, 1518 Clifton Rd, Atlanta, GA 30322

*Contributed equally

**S6-** Overview of Included Studies

| Author, Year | Study Design | Country | Location | Setting | Study Population(s) | Total Sample Size | Theory | COM-B codes | MMAT (qual final score) |
|---|---|---|---|---|---|---|---|---|---|
| Aberese-Ako 2023 | Qualitative | Ghana | Rural and Urban | Domestic - Household | Adults (Women and Men) | 64 | Not reported | O-Physical, O-Social, M-Reflective, and M-Automatic | 5 |
| Affleck 2012 | Mixed Methods | Bangladesh | Rural | Domestic - Household | Adults (Women only) | 34 | Not reported | O-Physical, M-Reflective, and M-Automatic | 5 |
| Afolabi 2022 | Qualitative | Senegal | Rural and Urban | Domestic - Households | Caregivers of children | 100 | Not reported | O-Physical, O-Social, M-Reflective, and M-Automatic | 5 |
| Akter 2014 | Qualitative | Bangladesh | Rural | Domestic - Household | Adults (Women only) | 144 | Not reported | C-Physical, C-Psychological, O-Physical, M-Reflective, and M-Automatic | 5 |
| Akter 2022 | Mixed Methods | Bangladesh | Rural | Domestic - Household | Adults (Women and Men) | 746 | Not reported | O-Physical | 5 |
| Al-Naggar 2013 | Qualitative | Malaysia | Not Reported | Institutions - Universities | Adults (Women and Men); Other: Medical University Students | 40 | Not reported | O-Physical, M-Reflective, and M-Automatic | 5 |
| Arendt 2015 | Mixed Methods | USA | Not Reported | Institutions - Workplace | Food workers | 25 | Not reported | C-Psychological and M-Automatic | 5 |
| Ashraf 2017 | Mixed Methods | Bangladesh | Rural | Domestic – Household | Adults (Women and Men) | 385 | IBM-WASH | O-Physical and O-Social | 5 |
| Atuyambe 2011 | Mixed Methods | Uganda | 888 – Not Reported | Domestic - Household | General population | 397 | Not reported | O-Physical and M-Automatic | 5 |
| Azam 2022 | Mixed Methods | Pakistan | Not Reported | Domestic - Household | General population | 400 | Not reported | O-Physical, O-Social, M-Reflective, and M-Automatic | 5 |
| Babalobi 2013 | Mixed Methods | Nigeria | Urban | Public - Markets | Children (Girls and Boys) | Not Reported | Not reported | C-Psychological, O-Physical, and M-Reflective | 5 |
| Bauza 2021 | Mixed Methods | India | Rural | Domestic – Household | Adults (Women and Men) | 131 | Not reported | C-Psychological, O-Physical, and M-Automatic | 5 |
| Biran 2005 | Mixed Methods | Kyrgyzstan | Rural | Domestic – Household | General population | Not Reported | Not reported | C-Psychological, O-Physical, and M-Automatic | 5 |
| Biran 2012 | Mixed Methods | Thailand, Ethiopia and Kenya | Other: refugee camps | Other: refugee camps | General population | 1127 | Not reported | O-Physical | 5 |
| Biswas 2017 | Mixed Methods | Bangladesh | Rural | Domestic – Household | General population | 80 | Not reported | O-Physical, O-Social, M-Reflective, and M-Automatic | 5 |
| Blum 2019 | Qualitative | Congo | Not Reported | Internally displaced person camps | General population | 18 | IBM-WASH | O-Physical, O-Social, M-Reflective, and M-Automatic | 5 |
| Chidziwisano 2019 | Mixed Methods | Malawi | Rural | Domestic – Household | Children (Girls and Boys) and Caregivers | 323 | RANAS | O-Physical, O-Social, and M-Automatic | 5 |
| Claude 2020 | Mixed Methods | Congo | Rural | Internally displaced person camps | General population | 307 | Not reported | O-Physical | 5 |
| Curtis 2003 | Mixed Methods | UK | Not Reported | Domestic – Household | Adults (Women and Men) | 33 | Not reported | M-Reflective, and M-Automatic | 5 |
| Dearden 2002 | Qualitative | Vietnam | Rural | Domestic – Household | Children (Girls and Boys) | 100 | Elicitation procedure | O-Physical, O-Social, M-Reflective, and M-Automatic | 5 |

**S6-** Overview of Included Studies

| | | | | | | | | | |
|---|---|---|---|---|---|---|---|---|---|
| Demberere 2016 | Mixed Methods | Zimbabwe | Rural | Domestic – Household | Mother-child dyads | Not Reported | Not reported | O-Physical and M-Automatic | 5 |
| Devkota 2020 | Qualitative | Nepal | Not Reported | Institutions - Schools | Adults (Women and Men); Children (Girls and Boys) | 50 | Not reported | O-Physical and M-Automatic | 2 |
| Didier 2021 | Mixed Methods | France, Norway, Portugal, Romania, UK | Rural and Urban | Domestic – Household | Adults (Women and Men) | Not Reported | Not reported | C-Psychological, O-Physical, M-Reflective, and M-Automatic | 5 |
| Grant 2023 | Mixed Methods | Bangladesh | Not Reported | Domestic – Household | General population | 1762 | Not reported | O-Physical | 5 |
| Green 2005 | Qualitative | USA | Not Reported | Institutions - Workplace | Food workers | Not Reported | Not reported | C-Psychological, O-Physical, O-Social, and M-Reflective | 5 |
| Greenwell 2013 | Qualitative | Fiji | Peri-urban | Domestic – Household | General population | 27 | Not reported | O-Physical, M-Reflective, and M-Automatic | 5 |
| Harrison 2019 | Qualitative | Uganda | Rural | Domestic – Household | Adults (Women only) | 55 | Not reported | O-Social and M-Reflective | 5 |
| Herbst 2009 | Mixed Methods | Vietnam | Peri-urban | Domestic – Household | General population | 135 | Not reported | M-Reflective | 5 |
| Hoque 2023 | Mixed Methods | Bangladesh | Rural | Domestic – Household | Adults (Women and Men) | 242 | Not reported | C-Psychological, O-Physical, and O-Social | 5 |
| Jackson 2021 | Qualitative | USA | Not Reported | Institutions - Schools | Adults (Women and Men) | 11 | Not reported | O-Physical, O-Social, M-Reflective, and M-Automatic | 5 |
| Kalam 2021 | Mixed Methods | Bangladesh | Urban | Domestic – Household | General population | 90 | RANAS; Behavior Centered Design/Evo-Eco Model; Health Belief Model | O-Physical | 5 |
| Kalumbi 2020 | Mixed Methods | Malawi | Not Reported | Domestic – Household | General population | 265 | Not reported | O-Physical, O-Social, M-Reflective, and M-Automatic | 5 |
| Kumar 2018 | Mixed Methods | India | Urban | Institutions - Schools | Children (Girls and Boys) | 194 | Not reported | C-Psychological, O-Physical, O-Social, and M-Automatic | 5 |
| La Con 2017 | Mixed Methods | Kenya | Rural | Institutions - Schools | Children (Girls and Boys) | Not Reported | Not reported | C-Psychological, O-Physical, and M-Reflective | 5 |
| Lando 2018 | Mixed Methods | USA | Not Reported | Domestic - Households; Institutions - Schools | Adults (Women and Men) | Not Reported | Not reported | M-Automatic | 5 |
| Lanfer 2021 | Qualitative | Sierra Leone | Rural and Urban | Domestic – Household | General population | 56 | IBM-WASH | O-Physical, O-Social, and M-Reflective | 5 |
| Langford 2013 | Mixed Methods | Nepal | Peri-urban | Domestic – Household | Adults (Women only) | Not Reported | Not reported | C-Psychological, O-Physical, O-Social, M-Reflective, and M-Automatic | 5 |
| Lohiniva 2007 | Qualitative | Egypt | Rural | Domestic - Households; Public - Markets | Adults (Women only) | Not Reported | Not reported | O-Physical, M-Reflective, and M-Automatic | 5 |

**S6-** Overview of Included Studies

| | | | | | | | | | |
|---|---|---|---|---|---|---|---|---|---|
| Mbakaya 2019 | Qualitative | Malawi | Not Reported | Institutions - Schools | Children (Girls and Boys) | Not Reported | Not reported | C-Psychological, O-Physical, O-Social, and M-Reflective | 5 |
| Melaku 2023 | Mixed Methods | Ethiopia | Not Reported | Institutions - Schools | Adults (Women and Men); Children (Girls and Boys) | Not Reported | Not reported | O-Physical | 5 |
| Mezaache 2021 | Mixed Methods | France | Urban | Other: harm reduction programs | Other: people who inject drugs | 59 | Not reported | M-Reflective and M-Automatic | 2 |
| Mitchell 2021 | Mixed Methods | Australia | Not Reported | Domestic – Household | Adults (Women and Men) | 426 | Not reported | C-Physical, C-Psychological, and M-Automatic | 5 |
| Mohamed 2022 | Qualitative | Tanzania | Urban | Domestic – Household | General population | 102 | Not reported | C-Physical, O-Physical, O-Social, and M-Reflective | 3 |
| Mshida 2020 | Mixed Methods | Tanzania | Rural | Domestic - Households; Public - Markets | General population | 3547 | Not reported | M-Reflective | 5 |
| Neetu 2013 | Qualitative | India | Not Reported | Public - Markets; Institutions - Workplace | Food workers | 67 | Not reported | O-Physical, M-Reflective, and M-Automatic | 5 |
| Nizame 2013 | Mixed Methods | Bangladesh | Rural | Domestic – Household | General population | 350 | COM-B | O-Physical, M-Reflective, and M-Automatic | 5 |
| Nizame 2016 | Qualitative | Bangladesh | Rural | Domestic – Household | General population | 24 | IBM-WASH; PRECEDE/PROCEED | O-Physical, M-Reflective, and M-Automatic | 5 |
| Nizame 2019 | Mixed Methods | Bangladesh | Rural and Urban | Public - Markets | General population; Food workers | 600 | IBM-WASH | C-Physical, O-Physical, and M-Reflective | 5 |
| Norrie 2022 | Qualitative | England | Not Reported | Domestic – Household | Adults (Women and Men) | 41 | Not reported | M-Reflective | 5 |
| Ntakirutimana 2021 | Mixed Methods | Rwanda | Not Reported | Domestic – Household | Adults (Women only) and Children (Girls and Boys) | Not Reported | Not reported | O-Physical | 5 |
| Ogutu 2022 | Qualitative | Kenya | Rural and Peri-urban | Domestic – Household | Adults (Women and Men) | 65 | COM-B | C-Psychological, O-Physical, M-Reflective, and M-Automatic | 5 |
| Okello 2019 | Qualitative | Tanzania | Rural and Urban | Institutions - Schools | Children (Girls and Boys) | 16 adults, 100 children | Behavior Centered Design/Evo-Eco Model; COM-B | C-Psychological, O-Physical, M-Reflective, and M-Automatic | 5 |
| Parveen 2018 | Qualitative | Bangladesh | Rural | Domestic – Household | Adults (Women only) | 64 | Theory of Planned Behavior; Health Belief Model | O-Physical, O-Social, M-Reflective, and M-Automatic | 5 |
| Pragle 2007 | Qualitative | USA | Rural and Urban | Institutions - Workplace | Food workers | 18 | Not reported | O-Physical, O-Social, M-Reflective, and M-Automatic | 5 |
| Rahman 2017 | Qualitative | Bangladesh | Urban | Domestic – Household | General population | 61 | Cultural Consensus Theory, Prototype Theory | C-Psychological. O-Social, and M-Reflective | 5 |

| | | | | | | | | | |
|---|---|---|---|---|---|---|---|---|---|
| Randle 2013 | Mixed Methods | England | Not Reported | Institutions - Schools; Other: healthcare setting | Children (Girls and Boys) | Not Reported | Not reported | M-Reflective and M-Automatic | 5 |
| Rauyajin 1994 | Qualitative | Thailand | Not Reported | Domestic – Household | Mother-child dyads | 12 | Health Belief Model; Model of families use of health service | M-Reflective and M-Automatic | 5 |
| Sagan 2019 | Mixed Methods | Nepal, Pakistan, and the Philippines | Rural and Urban | Domestic – Household | Adults (Women only) | Not Reported | Behavior Centered Design/Evo-Eco Model | O-Physical, O-Social, M-Reflective, and M-Automatic | 5 |
| Schmidt 2009 | Qualitative | UK | Urban | Institutions - Schools | Adults (Women and Men); Children (Girls and Boys) | Not Reported | Not reported | C-Psychological, O-Social, M-Reflective, and M-Automatic | 5 |
| Scott 2007 | Qualitative | Ghana | Rural and Urban | Domestic – Household | General population | 448 | Health Belief Model | O-Physical, O-Social, M-Reflective, and M-Automatic | 5 |
| Sebong 2021 | Qualitative | Indonesia | Urban | Institutions - Universities | Adults (Women and Men) | 23 | Not reported | C-Psychological, O-Physical, O-Social, and M-Reflective | 5 |
| Sedekia 2022 | Qualitative | Tanzania | Rural | Domestic - Households; Institutions - Schools | Children (Girls and Boys) | 20 | COM-B | C-Psychological and M-Automatic | 5 |
| Simiyu 2020 | Qualitative | Kenya | Peri-urban | Domestic – Household | Adults (Women and Men); Children (Girls and Boys) | 40 adults, 127 children | Behavior Centered Design/Evo-Eco Model | M-Reflective | 5 |
| Steenkamp 2022 | Qualitative | South Africa | Urban | Other: Early childhood development (ECD) centers | Adults (Women and Men); Children (Girls and Boys) | 105 | Not reported | O-Physical and M-Reflective | 5 |
| Steiner-Asiedu 2011 | Mixed Methods | Ghana | Not Reported | Domestic - Households; Institutions - Schools | Children (Girls and Boys) | 295 | Not reported | O-Physical and M-Reflective | 5 |
| Sultana 2018 | Qualitative | Bangladesh | Urban | Domestic - Households; Institutions - Schools | General population | Not Reported | IBM-WASH | O-Physical, O-Social, M-Reflective, and M-Automatic | 5 |
| Thaivalappil 2022 | Qualitative | Canada | Not Reported | Domestic – Household | Adults (Women and Men) | 37 | Theoretical Domains Framework | O-Social, M-Reflective, and M-Automatic | 5 |
| Thorseth 2021 | Mixed Methods | Ethiopia | Not Reported | Internally displaced person camps | General population | Not Reported | Not reported | O-Physical, M-Reflective, and M-Automatic | 5 |
| Tibbels 2022 | Qualitative | Coˆte d'Ivoire | Urban | Domestic – Household | General population | 152 | Not reported | O-Physical, O-Social, M-Reflective, and M-Automatic | 5 |
| Torres-Slimming 2019 | Mixed Methods | Peru | Rural | Domestic – Household | General population | 64 | Not reported | O-Physical, O-Social, and M-Reflective | 5 |

**S6-** Overview of Included Studies

| | | | | | | | | | |
|---|---|---|---|---|---|---|---|---|---|
| Ward 2022 | Mixed Methods | Nigeria, Burkina Faso, and Chad | Urban and Rural | Domestic – Household | Adults (Women and Men) | Not Reported | Donabedian's Model for Assessing Quality of Care | O-Social, M-Reflective, and M-Automatic | 5 |
| Watson 2020 | Qualitative | Iraq | Not Reported | Internally displaced person camps | Children (Girls and Boys) | 72 | IBM-WASH; Behavior Centered Design/Evo-Eco Model | O-Physical, O-Social, and M-Reflective | 5 |
| White 2022 A | Qualitative | Iraq | Not Reported | Domestic - Households; Internally displaced person camps | General population | 159 | Behavior Centered Design/Evo-Eco Model | O-Physical, O-Social, M-Reflective, and M-Automatic | 5 |
| White 2022 B | Qualitative | Congo | Rural and Urban | Domestic - Households; Internally displaced person camps | General population | 104 | Behavior Centered Design/Evo-Eco Model | O-Physical, M-Reflective, and M-Automatic | 4 |
| Wu 2019 | Mixed Methods | China | Urban | Public - Parks, squares, or other public outdoor spaces | Adults (Women and Men) | 300 | Not reported | O-Physical, M-Reflective, and M-Automatic | 5 |
| Xuan 2013 | Mixed Methods | Vietnam | Rural | Institutions - Schools | Children (Girls and Boys) | 343 | Not reported | O-Physical | 5 |
| Yallew 2012 | Mixed Methods | Ethiopia | Not Reported | Domestic – Household | Adults (Women and Men) | 258 | Not reported | O-Physical | 5 |
| Yardley 2011 | Mixed Methods | UK | Not Reported | Domestic – Household | General population | 129 | Theory of Planned Behavior | O-Physical, O-Social, M-Reflective, and M-Automatic | 5 |
| Yeasmin 2021 | Qualitative | Bangladesh | Rural | Domestic – Household | Adults (Women and Men) | 89 | IBM-WASH | O-Physical and O-Social | 3 |
| Zangana 2020 | Mixed Methods | Iraq | Other: Camps | Domestic - Households; Internally displaced person camps | General population | 180 | Not reported | C-Psychological, O-Physical, O-Social, M-Reflective, and M-Automatic | 5 |
