## Supplementary material for "Behavioural factors influencing hand hygiene practices across domestic, institutional, and public community settings: A systematic review": S7

**S7 –** Quality Appraisal of all Included Articles Using the Mixed Methods Appraisal Tool

1 Hubert Department of Global Health, Rollins School of Public Health, Emory University, Atlanta, GA, USA; (BAC); (DMS); (SKP)
2 Gangarosa Department of Environmental Health, Rollins School of Public Health, Emory University, Atlanta, GA, USA; (MCF); (MW) (JSS); (LAO); Kennedy Files (KF); Erin LaFon (EL)
3 Woodruff Health Sciences Center Library, Emory University, Atlanta, GA, USA; (HR)
4 Department of Disease Control, London School of Hygiene and Tropical Medicine, London, UK; (OC)
5 Water, Sanitation, Hygiene and Health Unit, World Health Organization, Geneva, Switzerland; (JEM)
Emory University, Rollins School of Public Health, 1518 Clifton Rd, Atlanta, GA 30322

*Contributed equally

**S7 –** Quality Appraisal of all Included Articles Using the Mixed Methods Appraisal Tool

**Quality Appraisal of all Included Articles Using the Mixed Methods Appraisal Tool**

| Study | Final Score | Qualitative Score | Mixed Methods Score | Criteria from the Mixed Methods Appraisal Tool[1] | | | | | | | | | |
|---|---|---|---|---|---|---|---|---|---|---|---|---|---|
| | | | | KEY<br>Individual criteria scores can be either 0 (did not meet criteria) or 1 (met criteria).<br><br>Qualitative and quantitative studies were assessed using the five-criteria questionnaire. Mixed methods studies were assessed using the relevant independent questionnaires for qualitative and quantitative work and a five criteria questionnaire for mixed methods; the lowest of the three scores was used as the quality score. Possible scores are 0–5 across study types (5 is the best).<br><br>Final scores represent only the qualitative components of the MMAT assessment because only qualitative data were extracted from the mixed methods studies included in the review.<br><br>† Indicates that the MMAT was deemed inappropriate for quality appraisal of the article. | | | | | | | | | |
| | | | | 1.1 | 1.2 | 1.3 | 1.4 | 1.5 | 5.1 | 5.2 | 5.3 | 5.4 | 5.5 |
| Aberese-Ako 2023 | 5 | 5 | | 1 | 1 | 1 | 1 | 1 | | | | | |
| Affleck 2012 | 5 | 5 | 5 | 1 | 1 | 1 | 1 | 1 | 1 | 1 | 1 | 1 | 1 |
| Afolabi 2022 | 5 | 5 | | 1 | 1 | 1 | 1 | 1 | | | | | |
| Akter 2014 | 5 | 5 | | 1 | 1 | 1 | 1 | 1 | | | | | |
| Akter 2022 | 5 | 5 | 5 | 1 | 1 | 1 | 1 | 1 | 1 | 1 | 1 | 1 | 1 |
| Al-Naggar 2013 | 5 | 5 | | 1 | 1 | 1 | 1 | 1 | | | | | |
| Arendt 2015 | 5 | 5 | 5 | 1 | 1 | 1 | 1 | 1 | 1 | 1 | 1 | 1 | 1 |
| Ashraf 2017 | 5 | 5 | 5 | 1 | 1 | 1 | 1 | 1 | 1 | 1 | 1 | 1 | 1 |
| Atuyambe 2011 | 5 | 5 | 3 | 1 | 1 | 1 | 1 | 1 | 1 | 1 | 0 | 0 | 1 |
| Azam 2022 | 5 | 5 | 0 | 1 | 1 | 1 | 1 | 1 | 0 | 0 | 0 | 0 | 0 |
| Babalobi 2013 | 5 | 5 | 5 | 1 | 1 | 1 | 1 | 1 | 1 | 1 | 1 | 1 | 1 |
| Bauza 2021 | 5 | 5 | 4 | 1 | 1 | 1 | 1 | 1 | 1 | 1 | 1 | 0 | 1 |

**S7 –** Quality Appraisal of all Included Articles Using the Mixed Methods Appraisal Tool

| | | | | | | | | | | | | | |
|---|---|---|---|---|---|---|---|---|---|---|---|---|---|
| Biran 2005 | 5 | 5 | 5 | 1 | 1 | 1 | 1 | 1 | 1 | 1 | 1 | 1 | 1 |
| Biran 2012 | 5 | 5 | 5 | 1 | 1 | 1 | 1 | 1 | 1 | 1 | 1 | 1 | 1 |
| Biswas 2017 | 5 | 5 | 5 | 1 | 1 | 1 | 1 | 1 | 1 | 1 | 1 | 1 | 1 |
| Blum 2019 | 5 | 5 | | 1 | 1 | 1 | 1 | 1 | | | | | |
| Chidziwisano 2019 | 5 | 5 | 5 | 1 | 1 | 1 | 1 | 1 | 1 | 1 | 1 | 1 | 1 |
| Claude 2020 | 5 | 5 | 4 | 1 | 1 | 1 | 1 | 1 | 1 | 1 | 1 | 0 | 1 |
| Curtis 2003 | 5 | 5 | 2 | 1 | 1 | 1 | 1 | 1 | 1 | 1 | 0 | 0 | 0 |
| Dearden 2002 | 5 | 5 | | 1 | 1 | 1 | 1 | 1 | | | | | |
| Demberere 2016 | 5 | 5 | 3 | 1 | 1 | 1 | 1 | 1 | 1 | 1 | 1 | 0 | 0 |
| Devkota 2020 | 2 | 2 | | 1 | 1 | 0 | 0 | 0 | | | | | |
| Didier 2021 | 5 | 5 | 5 | 1 | 1 | 1 | 1 | 1 | 1 | 1 | 1 | 1 | 1 |
| Grant 2023 | 5 | 5 | 5 | 1 | 1 | 1 | 1 | 1 | 1 | 1 | 1 | 1 | 1 |
| Green 2005 | 5 | 5 | | 1 | 1 | 1 | 1 | 1 | | | | | |
| Greenwell 2013 | 5 | 5 | | 1 | 1 | 1 | 1 | 1 | | | | | |
| Harrison 2019 | 5 | 5 | 4 | 1 | 1 | 1 | 1 | 1 | 1 | 1 | 1 | 0 | 1 |
| Herbst 2009 | 5 | 5 | 5 | 1 | 1 | 1 | 1 | 1 | 1 | 1 | 1 | 1 | 1 |
| Hoque 2023 | 5 | 5 | 5 | 1 | 1 | 1 | 1 | 1 | 1 | 1 | 1 | 1 | 1 |
| Jackson 2021 | 5 | 5 | | 1 | 1 | 1 | 1 | 1 | | | | | |
| Kalam 2021 | 5 | 5 | 4 | 1 | 1 | 1 | 1 | 1 | 1 | 1 | 1 | 0 | 1 |
| Kalumbi 2020 | 5 | 5 | 5 | 1 | 1 | 1 | 1 | 1 | 1 | 1 | 1 | 1 | 1 |
| Kumar 2018 | 5 | 5 | 5 | 1 | 1 | 1 | 1 | 1 | 1 | 1 | 1 | 1 | 1 |
| La Con 2017 | 5 | 5 | 3 | 1 | 1 | 1 | 1 | 1 | 1 | 1 | 1 | 0 | 0 |
| Lando 2018 | 5 | 5 | 5 | 1 | 1 | 1 | 1 | 1 | 1 | 1 | 1 | 1 | 1 |
| Lanfer 2021 | 5 | 5 | | 1 | 1 | 1 | 1 | 1 | | | | | |
| Langford 2013 | 5 | 5 | 5 | 1 | 1 | 1 | 1 | 1 | 1 | 1 | 1 | 1 | 1 |
| Lohiniva 2007 | 5 | 5 | | 1 | 1 | 1 | 1 | 1 | | | | | |
| Mbakaya 2019 | 5 | 5 | | 1 | 1 | 1 | 1 | 1 | | | | | |
| Melaku 2023 | 5 | 5 | 5 | 1 | 1 | 1 | 1 | 1 | 1 | 1 | 1 | 1 | 1 |

**S7 –** Quality Appraisal of all Included Articles Using the Mixed Methods Appraisal Tool

| | | | | | | | | | | | | | |
|---|---|---|---|---|---|---|---|---|---|---|---|---|---|
| Mezaache 2021 | 2 | 2 | 5 | 0 | 0 | 1 | 1 | 0 | 1 | 1 | 1 | 1 | 1 |
| Mitchell 2021 | 5 | 5 | 5 | 1 | 1 | 1 | 1 | 1 | 1 | 1 | 1 | 1 | 1 |
| Mohamed 2022 | 3 | 3 | | 1 | 0 | 0 | 1 | 1 | | | | | |
| Mshida 2020 | 5 | 5 | 5 | 1 | 1 | 1 | 1 | 1 | 1 | 1 | 1 | 1 | 1 |
| Neetu 2013 | 5 | 5 | | 1 | 1 | 1 | 1 | 1 | | | | | |
| Nizame 2013 | 5 | 5 | 5 | 1 | 1 | 1 | 1 | 1 | 1 | 1 | 1 | 1 | 1 |
| Nizame 2016 | 5 | 5 | | 1 | 1 | 1 | 1 | 1 | | | | | |
| Nizame 2019 | 5 | 5 | 5 | 1 | 1 | 1 | 1 | 1 | 1 | 1 | 1 | 1 | 1 |
| Norrie 2022 | 5 | 5 | | 1 | 1 | 1 | 1 | 1 | | | | | |
| Ntakirutimana 2021 | 5 | 5 | 5 | 1 | 1 | 1 | 1 | 1 | 1 | 1 | 1 | 1 | 1 |
| Ogutu 2022 | 5 | 5 | | 1 | 1 | 1 | 1 | 1 | | | | | |
| Okello 2019 | 5 | 5 | | 1 | 1 | 1 | 1 | 1 | | | | | |
| Parveen 2018 | 5 | 5 | | 1 | 1 | 1 | 1 | 1 | | | | | |
| Pragle 2007 | 5 | 5 | | 1 | 1 | 1 | 1 | 1 | | | | | |
| Rahman 2017 | 5 | 5 | | 1 | 1 | 1 | 1 | 1 | | | | | |
| Randle 2013 | 5 | 5 | 5 | 1 | 1 | 1 | 1 | 1 | 1 | 1 | 1 | 1 | 1 |
| Rauyajin 1994 | 5 | 5 | | 1 | 1 | 1 | 1 | 1 | | | | | |
| Sagan 2019 | 5 | 5 | 5 | 1 | 1 | 1 | 1 | 1 | 1 | 1 | 1 | 1 | 1 |
| Schmidt 2009 | 5 | 5 | | 1 | 1 | 1 | 1 | 1 | | | | | |
| Scott 2007 | 5 | 5 | | 1 | 1 | 1 | 1 | 1 | | | | | |
| Sebong 2021 | 5 | 5 | | 1 | 1 | 1 | 1 | 1 | | | | | |
| Sedekia 2022 | 5 | 5 | | 1 | 1 | 1 | 1 | 1 | | | | | |
| Simiyu 2020 | 5 | 5 | | 1 | 1 | 1 | 1 | 1 | | | | | |
| Steenkamp 2022 | 5 | 5 | | 1 | 1 | 1 | 1 | 1 | | | | | |
| Steiner-Asiedu 2011 | 5 | 5 | 5 | 1 | 1 | 1 | 1 | 1 | 1 | 1 | 1 | 1 | 1 |
| Sultana 2018 | 5 | 5 | | 1 | 1 | 1 | 1 | 1 | | | | | |
| Thaivalappil 2022 | 5 | 5 | | 1 | 1 | 1 | 1 | 1 | | | | | |

**S7 –** Quality Appraisal of all Included Articles Using the Mixed Methods Appraisal Tool

| | | | | | | | | | | | | | |
|---|---|---|---|---|---|---|---|---|---|---|---|---|---|
| Thorseth 2021 | 5 | 5 | 5 | 1 | 1 | 1 | 1 | 1 | 1 | 1 | 1 | 1 | 1 |
| Tibbels 2022 | 5 | 5 | | 1 | 1 | 1 | 1 | 1 | | | | | |
| Torres-Slimming 2019 | 5 | 5 | 1 | 1 | 1 | 1 | 1 | 1 | 0 | 0 | 1 | 0 | 0 |
| Ward 2022 | 5 | 5 | 5 | 1 | 1 | 1 | 1 | 1 | 1 | 1 | 1 | 1 | 1 |
| Watson 2020 | 5 | 5 | | 1 | 1 | 1 | 1 | 1 | | | | | |
| White 2022 | 5 | 5 | | 1 | 1 | 1 | 1 | 1 | | | | | |
| White 2022 | 4 | 4 | | 1 | 1 | 0 | 1 | 1 | | | | | |
| Wu 2019 | 5 | 5 | 4 | 1 | 1 | 1 | 1 | 1 | 1 | 1 | 1 | 0 | 1 |
| Xuan 2013 | 5 | 5 | 5 | 1 | 1 | 1 | 1 | 1 | 1 | 1 | 1 | 1 | 1 |
| Yallew 2012 | 5 | 5 | 5 | 1 | 1 | 1 | 1 | 1 | 1 | 1 | 1 | 1 | 1 |
| Yardley 2011 | 5 | 5 | 4 | 1 | 1 | 1 | 1 | 1 | 1 | 1 | 1 | 0 | 1 |
| Yeasmin 2021 | 3 | 3 | | 0 | 0 | 1 | 1 | 1 | | | | | |
| Zangana 2020 | 5 | 5 | 1 | 1 | 1 | 1 | 1 | 1 | 1 | 0 | 0 | 0 | 0 |

[1]Hong, Q.N., Pluye, P., et al. Mixed Methods Appraisal Tool (MMAT) Version 2018 User Guide. McGill Department of Family Medicine. 2018.

**Criteria from the MMAT:**

1.1 Is the qualitative approach appropriate to answer the research question?
1.2 Are the qualitative data collection methods adequate to address the research question?
1.3 Are the findings adequately derived from the data?
1.4 Is the interpretation of results sufficiently substantiated by data?
1.5 Is there coherence between qualitative data sources, collection, analysis and interpretation?
5.1 Is there an adequate rationale for using a mixed methods design to address the research question?
5.2 Are the different components of the study effectively integrated to answer the research question?
5.3 Are the outputs of the integration of qualitative and quantitative components adequately interpreted?
5.4 Are divergences and inconsistencies between quantitative and qualitative results adequately addressed?
5.5 Do the different components of the study adhere to the quality criteria of each tradition of the methods involved?
