## Supplementary material for "Behavioural factors influencing hand hygiene practices across domestic, institutional, and public community settings: A systematic review": S9

**S9.** Thematic analysis of barriers and enablers to hand hygiene practices in community settings categorized using the COM-B framework.

### Behavioural factors influencing hand hygiene practices across domestic, institutional, and public community settings: A systematic review

Bethany A. Caruso[1], Jedidiah S. Snyder[2], Lilly A. O'Brien[2], Erin LaFon[2] , Kennedy Files[2], Dewan Muhammad Shoaib[1], Sridevi K. Prasad[1] , Hannah Rogers[3] , Oliver Cumming[4,5], Joanna Esteves Mills[5], Bruce Gordon [5], Marlene K. Wolfe[2*], Matthew C. Freeman[2*]

1 Hubert Department of Global Health, Rollins School of Public Health, Emory University, Atlanta, GA, USA; (BAC); (DMS); (SKP)
2 Gangarosa Department of Environmental Health, Rollins School of Public Health, Emory University, Atlanta, GA, USA; (MCF); (MW) (JSS); (LAO); Kennedy Files (KF); Erin LaFon (EL)
3 Woodruff Health Sciences Center Library, Emory University, Atlanta, GA, USA; (HR)
4 Department of Disease Control, London School of Hygiene and Tropical Medicine, London, UK; (OC)
5 Water, Sanitation, Hygiene and Health Unit, World Health Organization, Geneva, Switzerland; (JEM)
Emory University, Rollins School of Public Health, 1518 Clifton Rd, Atlanta, GA 30322

*Contributed equally.

**S9.** Thematic analysis of barriers and enablers to hand hygiene practices in community settings categorized using the COM-B framework.

Thematic analysis of barriers and enablers to hand hygiene practices in community settings categorized using the COM-B framework.

| COM-B Construct | Domestic Settings | | | Institutional Settings | | | Public Settings | | |
|---|---|---|---|---|---|---|---|---|---|
| | Thematic summary of barriers (-) and enablers (+) | Source(s) | Cer-QUAL | Thematic summary of barriers (-) and enablers (+) | Source(s) | Cer-QUAL | Thematic summary of barriers (-) and enablers (+) | Source(s) | Cer-QUAL |
| **Physical Capability** - Physical strength, skill or stamina | | | | | | | | | |
| **Ease/ difficulty of washing hands** | **(-) N**oted constrained physical ability to wash children's hands because number of kids made it difficult | (-) n=1<br>Mitchell 2021 | *Very low* | - | (-) n=0 | - | (-) Vendor moving around noted physical challenge of carrying the water and soap needed to wash hands | (-) n=1<br>Nizame 2019 | *Very low* |
| | **(+)** - | (+) n=0 | - | (+)- | (+) n=0 | - | (+)- | (+) n=0 | - |
| **Ease/ difficulty of collecting water** | **(-)** Physical challenges walking to water source and carrying water noted as reasons for not being able to wash hands | (-) n=2<br>Akter 2014<br>Mohamed 2022 | *Very low* | - | (-) n=0 | - | (-)- | (-) n=0 | - |
| | **(+)** - | (+) n=0 | - | (+)- | (+) n=0 | - | (+)- | (+) n=0 | - |
| **Psychological Capability** - Knowledge/psychological strength, skills or stamina | | | | | | | | | |
| **Action knowledge** | **(-)** Insufficient or wrong information about when and how to wash hands limited practice | (-) n=6<br>Akter 2014<br>Biran 2005<br>Didier 2021<br>Hoque 2023<br>Langford 2013<br>Mitchell 2021 | *Mod.* | **(-)** Insufficient or wrong information about when and how to wash hands limited practice | (-) n=3<br>Arendt 2015<br>Kumar 2018<br>Schmidt 2009 | *Very low* | **(-)** | (-) n=0 | - |
| | **(+)** Having practical knowledge about when and how to wash hands reported to influence practice | (+) n=6<br>Akter 2014<br>Bauza 2021<br>Ogutu 2022<br>Rahman 2017<br>Sedekia 2022<br>Zangana 2020 | *Mod.* | **(+)** Having practical knowledge about when and how to wash hands reported to influence practice | (+) n=6<br>Babalobi 2013<br>La Con 2017<br>Green 2005<br>Mbakaya 2019<br>Okello 2019<br>Sebong 2021 | *Mod.* | **(+)** - | (+) n=0 | - |

**S9.** Thematic analysis of barriers and enablers to hand hygiene practices in community settings categorized using the COM-B framework.

| **Physical opportunity -** Opportunities provided by the environment, such as time, location and resource | | | | | | | | | |
|---|---|---|---|---|---|---|---|---|---|
| **Soap/Sanitizer availability** | **(-)** Lack of soap available consistently and in an accessible location hinders handwashing<br><br>Lack of sanitizer limits hand hygiene. | (-) n=13<br>Afolabi 2022<br>Biswas 2017<br>Didier 2021<br>Greenwell 2013<br>Kalumbi 2020<br>Lanfer 2021<br>Langford 2013<br>Lohiniva 2007<br>Mohamed 2022<br>Sagan 2019<br>Parveen 2018<br>Sultana 2018<br>White 2022 B | *High* | **(-)** Lack of soap available hinders handwashing<br><br>Lack of sanitizer limits hand hygiene. | (-) n=7<br>Al-Naggar 2013<br>Devkota 2020<br>Kumar 2018<br>Melaku 2023<br>Okello 2019<br>Sebong 2021<br>Steenkamp 2022 | *Mod.* | **(-)** Lack of soap hinders handwashing | (-) n=6<br>Babalobi 2013<br>Biran 2012<br>Blum 2019<br>Claude 2020<br>Thorseth 2021<br>Wu 2019 | *Mod.* |
| | **(+)** Soap availability enables handwashing.<br><br>Sanitizer availability enables hygiene. | (+) n=6<br>Biran 2005<br>Biswas 2017<br>Kalam 2021<br>Mohamed 2022<br>Sultana 2018<br>Zangana 2020 | *Mod.* | **(+)** Having soap enables handwashing practices. | (+) n=2<br>Jackson 2021<br>Mbakaya 2019 | *Very low* | **(+)** Soap availability enables handwashing | (+) n=2<br>Neetu 2013<br>Thorseth 2021 | *Very low* |
| **Soap cost** | **(-)** Soap is cost prohibitive, limiting handwashing | (-) n=20<br>Aberese-Ako 2023<br>Affleck 2012<br>Afolabi 2022<br>Akter 2022<br>Azam 2022<br>Bauza 2021<br>Biran 2005<br>Chidziwisano 2019<br>Biswas 2017<br>Grant 2023<br>Hoque 2023<br>Lanfer 2021<br>Langford 2013<br>Mohamed 2022<br>Nizame 2016<br>Ogutu 2022<br>Scott 2007<br>Parveen 2018<br>White 2022 A | *High* | **(-)** Ensuring availability of soap is a challenge at school due to cost, limiting handwashing. | (-) n=1<br>Devkota 2020 | *Very low* | **(-)** Soap is cost prohibitive, limiting handwashing | (-) n=3<br>Blum 2019<br>Nizame 2019<br>Thorseth 2021 | *Low* |

**S9.** Thematic analysis of barriers and enablers to hand hygiene practices in community settings categorized using the COM-B framework.

| | | | | | | | | | |
|---|---|---|---|---|---|---|---|---|---|
| | | Yallew 2012 | | | | | | | |
| | **(+)** Soap not considered too expensive for handwashing; soapy water is inexpensie and enables handwashing | (+) n=3<br>Biswas 2017<br>Ogutu 2022<br>Ashraf 2017 | *Low* | (+) - | (+) n=0 | - | (+) - | (+) n=0 | - |
| **Water availability** | **(-)** Lack of water hinders handwashing | (-) n=8<br>Akter 2014<br>Atuyambe 2011<br>Biran 2005<br>Demberere 2016<br>Didier 2021<br>Greenwell 2013<br>Lanfer 2021<br>Sagan 2019 | *High* | **(-)** Lack of water hinders handwashing | (-) n=3<br>Xuan 2013<br>Melaku 2023<br>Okello 2019 | *Low* | **(-)** Lack of water hinders handwashing | (-) n=2<br>Babalobi 2013<br>Claude 2020 | *Very low* |
| | **(+)** Availability of water enables handwashing | (+) n=3<br>Dearden 2002<br>Kalam 2021<br>Ashraf 2017 | *Very low* | **(+)** Availability of water enables handwashing | (+) n=2<br>Okello 2019<br>Steenkamp 2022 | *Very low* | (+) - | (+) n=0 | - |
| **Water supply quantity** | **(-)** No, limited, or irregular water supply constrained handwashing practice | (-) n=7<br>Aberese-Ako 2023<br>Akter 2014<br>Atuyambe 2011<br>Langford 201<br>Lohiniva 2007<br>Scott 2007<br>White 2022 B | *High* | **(-)** No or irregular water supply constrained handwashing practice | (-) n=2<br>Al-Naggar 2013<br>Melaku 2023 | *Very low* | **(-)** - | (-) n=0 | - |
| | **(+)** - | (+) n=0 | - | (+)- | (+) n=0 | - | (+)- | (+) n=0 | - |
| **Water distance** | **(-)** Distance to water limits use hand hygiene or amount of water used for hand hygiene | (-) n=5<br>Dearden 2002<br>Demberere 2016<br>Grant 2023<br>Mohamed 2022<br>Parveen 2018 | *Mod.* | **(-)** Lack of water supply near the school limited practice | (-) n=1<br>Al-Naggar 2013 | *Very low* | **(-)** - | (-) n=0 | - |
| | **(+)** Water close by facilitates practice | (+) n=2<br>Dearden 2002<br>Biswas 2017 | *Very low* | (+)- | (+) n=0 | - | (+)- | (+) n=0 | - |
| **Water cost** | **(-)** Cost of water limits handwashing practice | (-) n=2<br>Mohamed 2022<br>Yallew 2012 | *Very Low* | **(-)** - | (-) n=0 | - | **(-)** - | (-) n=0 | - |
| | **(+)** - | (+) n=0 | - | (+)- | (+) n=0 | - | (+)- | (+) n=0 | - |

**S9.** Thematic analysis of barriers and enablers to hand hygiene practices in community settings categorized using the COM-B framework.

| | | | | | | | | | |
|---|---|---|---|---|---|---|---|---|---|
| **Household infrastructure** | **(-)** No or limited access to household handwashing infrastructure/facility hinders practice | (-) n=5<br>Atuyambe 2011<br>Biswas 2017<br>Didier 2021<br>Hoque 2023<br>Lohiniva 2007 | *High* | **(-)** - | (-) n=0 | | **(-)** - | (-) n=0 | |
| | **(+)** Access to household handwashing infrastructure/facility enables practice | (+) n=1<br>Biswas 2017 | *Very Low* | (+)- | (+) n=0 | - | (+)- | (+) n=0 | - |
| **Community infrastructure** | **(-)** - | (-) n=0 | - | **(-)** No or limited access to handwashing infrastructure/facility hinders practice | (-) n=6<br>La Con 2017<br>Green 2005<br>Kumar 2018<br>Scott 2007<br>Sebong 2021<br>Steiner-Asiedu 2011 | *High* | **(-)** No or limited access to handwashing infrastructure/ facility hinders practice<br>- | (-) n=3<br>Aberese-Ako 2023<br>Scott 2007<br>Yardley 2011 | *Low* |
| | (+)- | (+) n=0 | - | **(+)** Access to handwashing infrastructure/facility enables practice | (+) n=1<br>Jackson 2021 | *Very Low* | (+)- | (+) n=0 | - |
| **Hand hygiene station location** | **(-)** Inconvenient location of handwashing stations—including distance, lack of proximity to bathrooms or cooking area, or locked away at night to prevent theft—limited practice | (-) n=6<br>Akter 2022<br>Chidziwisano 2019<br>Hoque 2023<br>Nizame 2013<br>Ashraf 2017<br>Steiner-Asiedu 2011 | *High* | **(-)** Inconvenient location of handwashing stations limited practice | (-) n=4<br>Al-Naggar 2013<br>La Con 2017<br>Green 2005<br>Jackson 2021 | *Mod.* | **(-)** Inconvenient location of handwashing stations limited practice | (-) n=1<br>Watson 2020 | *Very low* |
| | **(+)** Convenient location of handwashing station enabled practice | (+) n=3<br>Biswas 2017<br>Ashraf 2017<br>Tibbels 2022 | *Low* | **(+)** Convenient location of handwashing station enabled practice | (+) n=3<br>Jackson 2021<br>Okello 2019<br>Pragle 2007 | *Low* | **(+)** Convenient location of handwashing station enabled practice | (+) n=1<br>Neetu 2013 | *Very low* |
| **Hand hygiene station design** | **(-)** Poor design hindered use. Features noted include: poor quality, inaccessibility, lack of running water. | (-) n=4<br>Hoque 2023<br>Mohamed 2022<br>White 2022 B<br>Zangana 2020 | *Mod.* | **(-)** -<br>- | (-) n=0 | - | **(-)** Station designs reported to be inappropriately designed for people with disabilities (out of reach, hard to access) | (-) n=1<br>Mohamed 2022 | *Very low* |
| | **(+)** Clean and beautiful design with features like mirrors, basins to catch water, noted to enable use | (+) n=2<br>White 2022 A<br>White 2022 B | *Very low* | (+)- | (+) n=0 | - | (+)- | (+) n=0 | - |

**S9.** Thematic analysis of barriers and enablers to hand hygiene practices in community settings categorized using the COM-B framework.

| | | | | | | | | | |
|---|---|---|---|---|---|---|---|---|---|
| **State of hand hygiene equipment** | **(-)** Hand hygiene stations provided by NGO broke easily and limited sustained behavior; See as 'poor design for poor people' | (-) n=1<br>White 2020 B | *Very low* | **(-)** Poor student handwashing practices attributed to non-functional facilities that were not well maintained or stocked | (-) n=1<br>Melaku 2023 | *Very low* | **(-)** Non-functional handwashing stations, limit practice<br>- | (-) n=1<br>Blum 2019 | *Very low* |
| | (+)- | (+) n=0 | - | **(+)** Student handwashing practices attributed to functional facilities that are well maintained in private school | (+) n=1<br>Melaku 2023 | *Very low* | (+)- | (+) n=0 | - |
| **Cleanliness of handwashing environment** | **(-)** Dirty handwashing area limited practice | (-) n=1<br>Zangana 2020 | *Very low* | **(-)** Dirty handwashing area limited practice | (-) n=1<br>Kumar 2018 | *Very low* | **(-)** Dirty handwashing area limited practice | (-) n=1<br>Wu 2019 | *Very low* |
| | **(+)** - | (+) n=0 | - | (+)- | (+) n=0 | - | (+)- | (+) n=0 | - |
| **Lighting of handwashing area** | **(-) -** | (-) n=0 | - | **(-) -** | (-) n=0 | - | **(-)** Insufficient lighting to allow safe handwashing at night | (-) n=1<br>Watson 2020 | *Very low* |
| | **(+)** - | (+) n=0 | - | (+)- | (+) n=0 | - | (+)- | (+) n=0 | - |
| **Damage of resources** | (-) General damages to resources constrained hygiene practices | (-) n=1<br>White 2022 A | *Very low* | **(-)** Broken or damaged facilities limited practice | (-) n=3<br>La Con 2017<br>Melaku 2023<br>Pragle 2007 | *Low* | **(-)** - | (-) n=0 | - |
| | **(+)** - | (+) n=0 | - | (+)- | (+) n=0 | - | (+)- | (+) n=0 | - |
| **Theft of resources** | **(-)** Theft of soap hinders handwashing | (-) n=4<br>Akter 2014<br>Biswas 2017<br>Ntakirutimana 2021<br>Sultana 2018 | *Low* | **(-) -** | (-) n=0 | | **(-) -** | (-) n=0 | - |
| | **(+)** - | (+) n=0 | - | (+)- | (+) n=0 | - | (+)- | (+) n=0 | - |
| **Resources available for purchase** | **(-)** Limited soap/sanitizer in markets or access to markets limited practice | (-) n=2<br>Yeasmin 2021<br>Torres-Slimming 2019 | *Very Low* | **(-)** - | (-) n=0 | - | **(-)** Limited access to markets for soap limited practice (IDPs) | (-) n=1<br>Thorseth 2021 | *Very Low* |
| | **(+)** - | (+) n=0 | - | (+)- | (+) n=0 | - | (+)- | (+) n=0 | - |

**S9.** Thematic analysis of barriers and enablers to hand hygiene practices in community settings categorized using the COM-B framework.

| | | | | | | | | | |
|---|---|---|---|---|---|---|---|---|---|
| **Social Opportunity -** Opportunities because of social factors, like cultural norms and social cues | | | | | | | | | |
| **Role model for behavior** | **(-)** Lack of role models noted as influencing hand hygiene behaviors. | (1) n= 1<br>Lanfer 2021 | *Very low* | **(-)** - | (-) n=0 | - | **(-)** - | (-) n=0 | - |
| | **(+)** Child handwashing influenced by parents modeling behavior | (+) n=4<br>Yeasmin 2021<br>Langford 2013<br>Sagan 2019<br>Ashraf 2017 | *Low* | (+)- | (+) n=0 | - | (+)- | (+) n=0 | - |
| **Social pressure** | **(-)** Specific note of there not being social expectation or a social norm to practice hand hygiene or to use soap when washing hands. | (-) n=2<br>Langford 2013<br>Parveen 2018 | *Very Low* | **(-)-** | (-) n=0 | - | (-) | (-) n=0 | - |
| | **(+)** Various forms of social pressure noted as influencing hand washing, including: knowing people are watching, wanting to do what others are doing, being directly told. | (+) n=5<br>Afolabi 2022<br>Harrison 2019<br>Langford 2013<br>Thaivalappil 2022<br>Tibbels 2022 | *Mod.* | **(+)** Being seen by others (i.e., customers, co-workers, or teachers) noted to influence hand hygiene behaviors. | (+) n=4<br>Green 2005<br>Mbakaya 2019<br>Pragle 2007<br>Schmidt 2009 | *Mod.* | (+) Social pressure to conform to rules noted to influence hand hygiene behaviors. | (+) n=0<br>Blum 2019 | *Very Low* |
| **Visits by authorities** | **(-)** - | (-) n=0 | - | **(-)** - | (-) n=0 | - | **(-)** - | (-) n=0 | - |
| | **(+)** Unannounced visits by health workers motivated individuals to wash their hands more frequently. | (+) n=1<br>Ward 2022 | *Very Low* | **(+)** Non-compliance with handwashing in dorms attributed to lack of enforcement/consequences for students. | (+) n=1<br>Sebong 2021 | *Very Low* | (+)- | (+) n=0 | - |
| **Social stigmatization/ status** | **(-)** Concern with being teased or seen as 'obsessive' influence people to not wash hands or reduce intensity. | (-) n=4<br>Azam 2022<br>Parveen 2018<br>White 2022 B<br>Yardley 2011 | *Low* | **(-)** Students indicated fear of being teased by peers as limiting hand washing. | (-) n=1<br>Kumar 2018 | *Very Low* | **(-)** - | (-) n=0 | - |
| | **(+)** Concern for affiliation with others, respect, avoiding shame, being a good mother, what people might say, personal image influenced hand washing behavior. | (+) n=7<br>Langford 2013<br>Sagan 2019<br>Scott 2007<br>Tibbels 2022<br>White 2022 A<br>White 2022 B<br>Zangana 2020 | *Mod.* | **(+)** Desire for respect from others and a desire to prevent negative social consequences motivated hand washing behavior. | (+) n=2<br>Green 2005<br>Jackson 2021 | *Very Low* | **(+)** Desire to enhance image, have friends, and not be avoided or teased influenced children to wash their hands. | (+) n=2<br>Blum 2019,<br>Watson 2020 | *Very Low* |

**S9.** Thematic analysis of barriers and enablers to hand hygiene practices in community settings categorized using the COM-B framework.

| | | | | | | | | | |
|---|---|---|---|---|---|---|---|---|---|
| **Informational support** | **(-)** People with disabilities noted they lacked adequate timely information to influence handwashing during COVID because materials/information channels did not meet their needs (e.g., person with poor sight not able to see information on posters well) | (-) n=1<br>Mohamed 2022 | *Very Low* | **(-)** - | (-) n=0 | - | **(-)** - | (-) n=0 | - |
| | **(+)** Public health messages on preventing COVID-19 noted to influence hand washing practices | (+) n=2<br>Afolabi 2022<br>Torres-Slimming 2019 | *Very Low* | (+)- | (+) n=0 | - | (+)- | (+) n=0 | - |
| **Tangible/ instrumental support** | **(-)**- | (-) n=0 | | **(-)** - | (-) n=0 | | **(-)** - | (-) n=0 | |
| | **(+)** Tangible support from others, whether goods (soap, water, handwashing station) or assistance (e.g., setting up handwashing system), noted to influence hand washing practice | (+) n=3<br>Biswas 2017<br>Parveen 2018<br>Sultana 2018 | *Low* | (+)- | (+) n=0 | - | (+)- | (+) n=0 | - |
| **Network support** | **(-)** Lack of network support from family, and lack of personal connection with others who could support/remind about handwashing noted to negatively impact hand washing behavior | (-) n=3<br>Rahman 2017<br>Parveen 2018<br>White 2022 A | Low | **(-)** | (-) n=0 | - | (-) | (-) n=0 | - |
| | **(+)** Support/reminders from friends, neighbors, 'society', fellow members of 'groups', and others in networks noted to influence handwashing behaviors. | (+) n=7<br>Dearden 2002<br>Langford 2013<br>Rahman 2017<br>Scott 2007<br>Parveen 2018<br>Thaivalappil 2022<br>White 2022 A | *Mod.* | **(+)** Hand washing noted to be influenced by parents, work supervisors | (+) n=2<br>Green 2005<br>Schmidt 2009 | *Very low* | (+) Hygiene promoters, schools noted to influence handwashing behaviors | (+) n=1<br>Watson 2020 | *Very low* |

**S9.** Thematic analysis of barriers and enablers to hand hygiene practices in community settings categorized using the COM-B framework.

| | | | | | | | | | |
|---|---|---|---|---|---|---|---|---|---|
| **Cultural practices and norms** | **(-)** Cultural practice for handwashing not strong, handwashing seen to not align with culture (e.g., use of sanitizer with alcohol as against religion) or cultural expectations hinder handwashing (e.g., restrictions on movement). | (-) n=6<br>Aberese-Ako 2023<br>Hoque 2023<br>Kalumbi 2020<br>Parveen 2018<br>Ward 2022<br>White 2022 B | *Mod.* | **(-)** - | (-) n=0 | - | **(-)** - | (-) n=0 | - |
| | **(+)** Cultural practice for handwashing strong, no taboo in culture against handwashing, and cultural expectations and practices reinforce handwashing | (+) n=4<br>Afolabi 2022<br>Chidziwisano 2019<br>Thaivalappil 2022<br>Zangana 2020 | *Low* | **(+)** Established a norm within the culture of the training facility that all who entered would wash/sanitize hands, which influenced handwashing among those there | (+) n=1<br>Jackson 2021 | *Very Low* | (+)- | (+) n=0 | - |
| **Reflective Motivation** - Reflective processes, such as making plans and evaluating things that have already happened | | | | | | | | | |
| **Perceived health Risk** | **(-)** Limited perceived risk to health limits handwashing practices | (-) n=4<br>Azam 2022<br>Parveen 2018<br>Tibbels 2022<br>Thaivalappil 2022 | *Low* | **(-)** Limited perceived health risk limits handwashing practices; Perception that health will be impacted by using available (dirty) facilities limits handwashing | (-) n=2<br>Al-Naggar 2013<br>Sebong 2021 | *Very Low* | **(-)** - | (-) n=0 | - |
| | **(+)** Perceived risk to health—whether personal or those they care for— motivates, increases, or maintains handwashing practices; Perception that handwashing is good for health motivates practice | (+) n=24<br>Aberese-Ako 2023<br>Affleck 2012<br>Afolabi 2022<br>Akter 2014<br>Curtis 2003<br>Dearden 2002<br>Biswas 2017<br>Didier 2021<br>Kalumbi 2020<br>Lanfer 2021<br>Langford 2013<br>Lohiniva 2007<br>Nizame 2013<br>Norrie 2022<br>Ogutu 2022<br>Rahman 2017<br>Scott 2007<br>Parveen 2018<br>Steiner-Asiedu 2011<br>Tibbels 2022<br>Torres-Slimming 2019 | *High* | **(+)** Perceived risk to health—whether personal or of others (customers)— motivates handwashing practices | (+) n=5<br>Mbakaya 2019<br>Okello 2019<br>Pragle 2007<br>Schmidt 2009<br>Sebong 2021 | *Mod.* | **(+)** Perceived risk to health motivates handwashing practices | (+) n=3<br>Babalobi 2013<br>Blum 2019<br>Watson 2020 | *Very Low* |

**S9.** Thematic analysis of barriers and enablers to hand hygiene practices in community settings categorized using the COM-B framework.

| | | | | | | | | | |
|---|---|---|---|---|---|---|---|---|---|
| | | Ward 2022<br>White 2022 B<br>Zangana 2020 | | | | | | | |
| **Time prioritization** | **(-)** Any handwashing and handwashing with soap limited by how much time it takes, the lack of convenience, or because other activities or responsibilities compete for their time and are a priority, particularly among mothers. | (-) n=11<br>Affleck 2012<br>Akter 2014<br>Curtis 2003<br>Dearden 2002<br>Langford 2013<br>Lohiniva 2007<br>Nizame 2016<br>Rauyajin 1994<br>Sagan 2019<br>Parveen 2018<br>Steiner-Asiedu 2011 | *High* | **(-)** Time pressure, being busy, other priorities, and competing interests limit handwashing | (-) n=7<br>Al-Naggar 2013<br>La Con 2017<br>Green 2005<br>Jackson 2021<br>Pragle 2007<br>Schmidt 2009<br>Steenkamp 2022 | *High.* | **(-)** Time pressure, being busy, other priorities, and competing interests limit handwashing at all or by guideline/with soap | (-) n=5<br>Blum 2019<br>Mezaache 2021<br>Neetu 2013<br>Nizame 2019<br>Wu 2019 | *Mod.* |
| | **(+)** Handwashing thought of as non- time consuming, reinforcing practice, particularly in regard to soapy water | (+) n=2<br>Dearden 2002<br>Biswas 201 | *Very Low* | (+)- | (+) n=0 | - | (+)- | (+) n=0 | - |
| **Water prioritization** | **(-)** The need to ration, minimize, or prioritize water for others uses limits hand washing | (-) n=4<br>Lohiniva 2007<br>Mshida 2020<br>Ogutu 2022<br>Thorseth 2021 | *Mod.* | **(-)** - | (-) n=0 | - | **(-)** - | (-) n=0 | - |
| | **(+)** - | (+) n=0 | - | (+)- | (+) n=0 | - | (+)- | (+) n=0 | - |
| **Willingness to practice handwashing** | **(-)** Practice of handwashing or using sanitizer limited (e.g., not done at all, or not done with soap) based on various considerations, including that: hand washing is not necessary given activity/ circumstances or because hands are not really dirty; resources required for handwashng are too great; handwashing is not effective; handwashing is irritating to hands | (-) n=7<br>Affleck 2012<br>Didier 2021<br>Herbst 2009<br>Greenwell 2013<br>Mohamed 2022<br>Rauyajin 1994<br>Parveen 2018 | *Mod.* | **(-)** Practice of handwashing limited because hands not considered 'dirty enough' or because of concern for handwashing making skin chapped | (-) n=2<br>Al-Naggar 2013<br>Green 2005 | *Very Low* | **(-)** Practice of handwashing limited because hands not visibly dirty<br><br>- | (-) n=2<br>Thorseth 2021<br>Wu 2019 | *Very Low* |
| | **(+)** Practice of handwashing justified for varied reasons, including: soap is less costly than medicines, specific practice performed requires it (e.g., exposure to dirt), it enhances appetite, | (+) n=6<br>Akter 2014<br>Dearden 2002<br>Harrison 2019<br>Herbst 2009<br>Rahman 2017<br>Parveen 2018 | *Mod.* | **(+)** Concern for customer health motivated hand washing practice | (+) n=1<br>Pragle 2007 | *Very Low* | (+)- | (+) n=0 | - |

**S9.** Thematic analysis of barriers and enablers to hand hygiene practices in community settings categorized using the COM-B framework.

| | | | | | | | | | |
|---|---|---|---|---|---|---|---|---|---|
| **Perceived ease/difficulty of washing hands** | (-) Handwashing hindered because it was thought to be difficult to do for 30 seconds, not easy, a struggle to do with all else going on in life (at IDP camp) or with all other prevention measures (COVID). Hand-gel viewed as ineffective at removing dirt. | (-) n= 5<br>Dearden 2002<br>Norrie 2022<br>Ward 2022<br>White 2022 A<br>Yardley 2011 | *Mod.* | **(-)** Challenges needing to retrieve soap from another location hindered handwashing | (-) n=1<br>Steenkamp 2022 | *Very Low* | **(-)** Complicated procedures, hassle of carrying products needed for hand hygiene, and concern about product packaging (specifically for MONO-RUB) hindered practice. | (-) n=3<br>Mezaache 2021<br>Nizame 2019<br>Wu 2019 | *Very Low* |
| | (+) Practice of handwashing perceived to be easy, which facilitated behavior; perceived ease of using sanitizer facilitated use, particularly when other modes were perceived difficult for handwashing; specific ease of soapy water facilitated behavior. | (+) n= 7<br>Biswas 2017<br>Langford 2013<br>Mohamed 2022<br>Simiyu 2020<br>Sultana 2018<br>Ward 2022<br>White 2022 A | *Mod.* | **(+)** Ease and novelty of product (Glo-Yo) enabled hand hygiene among kids | (+) n=1<br>Randle 2013 | *Very Low* | **(+)** Ease of use, portability, and ability to use without water or sink were specific reasons driving use of 'MONO-RUB' for hand washing. | (+) n=1<br>Mezaache 2021 | *Very Low* |
| **Influence of religion** | (-) Reflections that God determined health or that practicing handwashing could imply a lack of faith deterred handwashing practice | (-) n=3<br>Azam 2022<br>Sagan 2019<br>Tibbels 2022 | *Very Low* | **(-)** | (-) n=0 | - | **(-)** | (-) n=0 | - |
| | (+) Handwashing practiced because it is a part of faith | (+) n=1<br>Afolabi 2022 | *Very Low* | (+)- | (+) n=0 | - | (+)- | (+) n=0 | - |

**S9.** Thematic analysis of barriers and enablers to hand hygiene practices in community settings categorized using the COM-B framework.

| **Automatic Motivation -** Automatic processes, such as our desires, impulses and inhibitions | | | | | | | | | |
|---|---|---|---|---|---|---|---|---|---|
| **Internal motivation/ Habit** | **(-)** Handwashing (generally or with soap) hindered by lack of motivation, routine, or habit; 'laziness', negligence, or carelessness; forgetting; presence of habits that do not include handwashing; not feeling disgust | (-) n=17<br>Aberese-Ako 2023<br>Affleck 2012<br>Akter 2014<br>Atuyambe 2011<br>Azam 2022<br>Curtis 2003<br>Dearden 2002<br>Didier 2021<br>Kalumbi 2020<br>Langford 2013<br>Lohiniva 2007<br>Nizame 2013<br>Nizame 2016<br>Rauyajin 1994<br>Parveen 2018<br>Thaivalappil 2022<br>Tibbels 2022 | *High* | **(-)** Lack of motivation to wash hands, often described as laziness | (-) n=3<br>Al-Naggar 2013<br>Kumar 2018<br>Schmidt 2009 | *Low* | **(-)** Lack of habit, routine, or ability to remember to practice handwashing (generally or with soap specifically) and presence of habits or routines that do not include handwashing | (-) n=4<br>Blum 2019<br>Mezaache 2021<br>Thorseth 2021<br>Wu 2019 | *Low* |
| | **(+)** Handwashing behavior driven by routine, habit, or 'need', particularly after certain activities (e.g., toileting), feeling of disgust, | (+) n=19<br>Affleck 2012<br>Bauza 2021<br>Biran 2005<br>Chidziwisano 2019<br>Curtis 2003<br>Dearden 2002<br>Biswas 2017<br>Didier 2021<br>Greenwell 2013<br>Kalumbi 2020<br>Lando 2018<br>Langford 2013<br>Mitchell 2021<br>Sagan 2019<br>Sedekia 2022<br>Parveen 2018<br>Sultana 2018<br>Thaivalappil 2022<br>Yardley 2011 | *High* | **(+)** 'Motivation' and habit driving practice | (+) n=6<br>Arendt 2015<br>Devkota 2020<br>Jackson 2021<br>Neetu 2013<br>Okello 2019<br>Pragle 2007 | *Mod.* | (+)- | (+) n=0 | - |

**S9.** Thematic analysis of barriers and enablers to hand hygiene practices in community settings categorized using the COM-B framework.

| | | | | | | | | | |
|---|---|---|---|---|---|---|---|---|---|
| **Feeling of cleanliness** | **(-)** - | (-) n=0 | - | **(-)** Handwashing not motivated because kids 'like dirt' | (-) n=1<br>Schmidt 2009 | *Very low* | **(-)** - | (-) n=0 | - |
| | **(+)** Handwashing motivated by desire to be comfortable, clean, and fresh; to look good; to remove smells and visible dirt; | (+) n=9<br>Afolabi 2022<br>Biran 2005<br>Curtis 2003<br>Demberere 2016<br>Ogutu 2022<br>Scott 2007<br>Parveen 2018<br>White 2022 A<br>Zangana 2020 | *High* | **(+)** Handwashing motivated because of want for cleanliness | (+) n=1<br>Schmidt 2009 | *Very low* | **(+)** Handwashing motivated by preference to be clean, enjoyment of cleanliness, refreshing feeling | (+) n=4<br>Blum 2019<br>Mezaache 2021<br>Neetu 2013<br>Wu 2019 | *Low* |
| **Like/dislike of the handwash product (e.g., feeling, smell, package)** | **(-)** Practice hindered because disliked; Specific dislike of alcohol sanitizers' effect on skin hindered use | (-) n=2<br>Dearden 2002<br>Ward 2022 | *Very low* | **(-)** - | (-) n=0 | - | **(-)** Dislike of handwash solution/packaging hindered practice | (-) n=1<br>Mezaache 2021 | *Very low* |
| | **(+)** Pleasant smell of soap and thus hands encouraged practice | (+) n=2<br>Biran 2005<br>Langford 2013 | *Very low* | **(+)** Enjoyment of product (Glo-Yo) and smell of soap enabled use. | (+) n=2<br>Randle 2013<br>Schmidt 2009 | *Very low* | **(+)** Approval of feel and smell of soap enabled practice | (+) n=3<br>Blum 2019<br>Mezaache 2021<br>Wu 2019 | *Low* |
