## Supplementary material for "Behavioural factors influencing hand hygiene practices across domestic, institutional, and public community settings: A systematic review": S10

**S10**- CERQual Assesment

1 Hubert Department of Global Health, Rollins School of Public Health, Emory University, Atlanta, GA, USA; (BAC); (DMS); (SKP)
2 Gangarosa Department of Environmental Health, Rollins School of Public Health, Emory University, Atlanta, GA, USA; (MCF); (MW) (JSS); (LAO); Kennedy Files (KF); Erin LaFon (EL)
3 Woodruff Health Sciences Center Library, Emory University, Atlanta, GA, USA; (HR)
4 Department of Disease Control, London School of Hygiene and Tropical Medicine, London, UK; (OC)
5 Water, Sanitation, Hygiene and Health Unit, World Health Organization, Geneva, Switzerland; (JEM)
Emory University, Rollins School of Public Health, 1518 Clifton Rd, Atlanta, GA 30322

*Contributed equally

**S10**- CERQual Assesment

| Summary of review finding | Setting | Barrier/ Enabler | Studies contributing to finding | Mean MMAT Score for Theme | Methodological limitations (Informed by MMAT)* | Coherence* | Adequacy* | Relevance* | Overall CERQual assessment of confidence in the evidence** | Explanation of CERQual assessment |
|---|---|---|---|---|---|---|---|---|---|---|
| **CAPABILITY** | | | | | | | | | | |
| **Physical Capability** | | | | | | | | | | |
| **Ease/ difficulty of washing hands** | **Domestic** | Barrier | Mitchell 2021 | 5 | No concerns | Serious concerns | Serious concerns | Serious concerns | Very Low | Very small number of studies limiting coherence, adequacy and geographical/population relevance. |
| | | Enabler | x | x | x | x | x | x | x | x |
| | **Institutional** | Barrier | x | x | x | x | x | x | x | x |
| | | Enabler | x | x | x | x | x | x | x | x |
| | **Public** | Barrier | Nizame 2019 | 5 | No concerns | Serious concerns | Serious concerns | Serious concerns | Very Low | Very small number of studies limiting coherence, adequacy and geographical/population relevance. |
| | | Enabler | x | x | x | x | x | x | x | x |
| **Ease/ difficulty of collecting water** | **Domestic** | Barrier | Akter 2014 Mohamed 2022 | 4 | Minor concerns | Serious concerns | Serious concerns | Serious concerns | Very Low | Very small number of studies limiting coherence, adequacy and geographical/population relevance. |
| | | Enabler | x | x | x | x | x | x | x | x |
| | **Institutional** | Barrier | x | x | x | x | x | x | x | x |
| | | Enabler | x | x | x | x | x | x | x | x |
| | **Public** | Barrier | x | x | x | x | x | x | x | x |
| | | Enabler | x | x | x | x | x | x | x | x |

**S10**- CERQual Assesment

| Psychological Capability | | | | | | | | | | |
|---|---|---|---|---|---|---|---|---|---|---|
| Action Knowledge | Domestic | Barrier | Akter 2014<br>Biran 2005<br>Didier 2021<br>Hoque 2023<br>Langford 2013<br>Mitchell 2021 | 5 | No concerns | Moderate concerns | Moderate concerns | Serious concerns | Moderate | Number of studies limiting coherence, adequacy, and geographical/population relevance (half in Asia, none in Africa; limited urban). |
| | | Enabler | Akter 2014<br>Bauza 2021<br>Ogutu 2022<br>Rahman 2017<br>Sedekia 2022<br>Zangana 2020 | 5 | No concerns | Moderate concerns | Moderate concerns | Serious concerns | Moderate | Number of studies limiting coherence, adequacy, and geographical/population relevance (most Africa/Asia, none in HICs; limited urban). |
| | Institutional | Barrier | Arendt 2015<br>Kumar 2018<br>Schmidt 2009 | 5 | No concerns | Moderate concerns | Serious concerns | Serious concerns | Very Low | Very small number of studies limiting coherence, adequacy, and geographical/population relevance (most in HICs; most in school settings). |
| | | Enabler | Babalobi 2013<br>La Con 2017<br>Green 2005<br>Mbakaya 2019<br>Okello 2019<br>Sebong 2021 | 5 | No concerns | Moderate concerns | Serious concerns | Serious concerns | Moderate | Number of studies limiting coherence, adequacy, and geographical/population relevance (half in Africa, many regions missing; most in educaiton settings). |
| | Public | Barrier | x | x | x | x | x | x | x | x |
| | | Enabler | x | x | x | x | x | x | x | x |

**S10**- CERQual Assesment

| OPPORTUNITY | | | | | | | | | | |
|---|---|---|---|---|---|---|---|---|---|---|
| **Physical Opportunity** | | | | | | | | | | |
| **Soap/ Sanitizer availability** | **Domestic** | Barrier | Afolabi 2022<br>Biswas 2017<br>Didier 2021<br>Greenwell 2013<br>Kalumbi 2020<br>Lanfer 2021<br>Langford 2013<br>Lohiniva 2007<br>Mohamed 2022<br>Sagan 2019<br>Parveen 2018<br>Sultana 2018<br>White 2022 B | 4.7 | Very minor concerns | No concerns | No concerns | Very minor concerns | High | Overall, adequate number of studies supporting theme across multiple settings. Some limitation in geographic relevance (HIC limited; most Africa, Asia). |
| | | Enabler | Biran 2005<br>Biswas 2017<br>Kalam 2021<br>Mohamed 2022<br>Sultana 2018<br>Zangana 2020 | 4.7 | Very minor concerns | No concerns | Very minor concerns | Moderate concerns | Moderate | Limitation in geographic relevance (No HIC; most Africa, Asia). |
| | **Institutional** | Barrier | Al-Naggar 2013<br>Devkota 2020<br>Kumar 2018<br>Melaku 2023<br>Okello 2019<br>Sebong 2021<br>Steenkamp 2022 | 4.6 | Very minor concerns | No concerns | Very minor concerns | Moderate concerns | Moderate | Limitation in geographic relevance (No HIC; all education settings). |
| | | Enabler | Jackson 2021<br>Mbakaya 2019 | 5 | No concerns | Moderate concerns | Serious concerns | Serious concerns | Very Low | Very small number of studies limiting coherence, adequacy, and geographical/population relevance( 1 Us, 1 Africa, all schools). |

| | | | | | | | | | | |
|---|---|---|---|---|---|---|---|---|---|---|
| **Soap/ Sanitizer availability** | **Public** | Barrier | Babalobi 2013<br>Biran 2012<br>Blum 2019<br>Claude 2020<br>Thorseth 2021<br>Wu 2019 | 5 | No concerns | Moderate concerns | Minor concerns | Serious concerns | Moderate | Number of studies limiting adequacy and geographical/population relevance (all Africa; half IDP). |
| | | Enabler | Neetu 2013<br>Thorseth 2021 | 5 | No concerns | Moderate concerns | Serious concerns | Serious concerns | Very Low | Very limited number of studies limiting adequacy and geographical/population relevance (no HICs; one market,1 IDP). |
| **Soap cost** | **Domestic** | Barrier | Aberese-Ako 2023<br>Affleck 2012<br>Afolabi 2022<br>Akter 2022<br>Azam 2022<br>Bauza 2021<br>Biran 2005<br>Chidziwisano 2019<br>Biswas 2017<br>Grant 2023<br>Hoque 2023<br>Lanfer 2021<br>Langford 2013<br>Mohamed 2022<br>Nizame 2016<br>Ogutu 2022<br>Scott 2007<br>Parveen 2018<br>White 2022 A<br>Yallew 2012 | 4.9 | Very minor concerns | No concerns | No concerns | Minor concerns | High | Overall, adequate number of studies supporting theme across multiple settingsLimitation in geographic relevance (No HIC; most Africa, Asia). |
| | | Enabler | Biswas 2017<br>Ogutu 2022<br>Ashraf 2017 | 5 | No concerns | Minor concerns | Serious concerns | Serious concerns | Low | Number of studies limiting adequacy and geographical/population relevance (No HICs; all rural, per-urban). |

**S10**- CERQual Assesment

| | | | | | | | | | | |
|---|---|---|---|---|---|---|---|---|---|---|
| **Soap cost** | **Institutional** | Barrier | Devkota 2020 | 2 | Serious concerns | Moderate concerns | Serious concerns | Serious concerns | Very Low | Number of studies limiting adequacy and geographical/population relevance (only Nepal, only school). |
| | | Enabler | x | x | x | x | x | x | x | x |
| | **Public** | Barrier | Blum 2019<br>Nizame 2019<br>Thorseth 2021 | 5 | No concerns | Minor concerns | Serious concerns | Serious concerns | Low | Number of studies limiting adequacy and geographical/population relevance (no HICs; 1 markey; IDPs). |
| | | Enabler | x | x | x | x | x | x | x | x |
| **Water availability** | **Domestic** | Barrier | Akter 2014<br>Atuyambe 2011<br>Biran 2005<br>Demberere 2016<br>Didier 2021<br>Greenwell 2013<br>Lanfer 2021<br>Sagan 2019 | 5 | No concerns | No concerns | Very minor concerns | Minor concerns | High | Overall, adequate number of studies supporting theme across multiple settings. Limitation in geographic relevance (most Africa, Asia). |
| | | Enabler | Dearden 2002<br>Kalam 2021<br>Ashraf 2017 | 5 | No concerns | Moderate concerns | Serious concerns | Serious concerns | Very Low | Limitation in geographic relevance (No HIC; all Asia). |
| | **Institutional** | Barrier | Xuan 2013<br>Melaku 2023<br>Okello 2019 | 5 | No concerns | Minor concerns | Serious concerns | Serious concerns | Low | Limitation in geographic relevance (No HIC; all schools). |
| | | Enabler | Okello 2019<br>Steenkamp 2022 | 5 | No concerns | Moderate concerns | Serious concerns | Serious concerns | Very Low | Limitation in geographic relevance (all Africa; all schools). |
| | **Public** | Barrier | Babalobi 2013<br>Claude 2020 | 5 | No concerns | Moderate concerns | Serious concerns | Serious concerns | Very Low | Limitation in geographic relevance (all Africa; 1 market, 2 IDP). |
| | | Enabler | x | x | x | x | x | x | x | x |

**S10**- CERQual Assesment

| | | | | | | | | | |
|---|---|---|---|---|---|---|---|---|---|
| Water supply quantity | Domestic | Barrier | Aberese-Ako 2023<br>Akter 2014<br>Atuyambe 2011<br>Langford 2013<br>Lohiniva 2007<br>Scott 2007<br>White 2022 B | 4.9 | Very minor concerns | No concerns | Minor concerns | Moderate concerns | High | Limitation in geographic relevance (No HIC; majority Afirica). |
| | | Enabler | x | x | x | x | x | x | x | x |
| | Institutional | Barrier | Al-Naggar 2013<br>Melaku 2023 | 5 | No concerns | Moderate concerns | Serious concerns | Serious concerns | Very Low | Limitation in geographic relevance (No HIC; Education settings only). |
| | | Enabler | x | x | x | x | x | x | x | x |
| | Public | Barrier | x | x | x | x | x | x | x | x |
| | | Enabler | x | x | x | x | x | x | x | x |
| Water distance | Domestic | Barrier | Dearden 2002<br>Demberere 2016<br>Grant 2023<br>Mohamed 2022<br>Parveen 2018 | 4.6 | Very minor concerns | No concerns | Minor concerns | Moderate concerns | Moderate | Limitation in geographic relevance (No HIC; Africa/Asia only). |
| | | Enabler | Dearden 2002<br>Biswas 2017 | 5 | No concerns | Moderate concerns | Serious concerns | Serious concerns | Very Low | Limitation in geographic relevance (No HIC; Asia only; rural only). |
| | Institutional | Barrier | Al-Naggar 2013 | 5 | No concerns | Moderate concerns | Serious concerns | Serious concerns | Very Low | Limitation in geographic relevance (No HIC; Asia only; University only). |
| | | Enabler | x | x | x | x | x | x | x | x |
| | Public | Barrier | x | x | x | x | x | x | x | x |
| | | Enabler | x | x | x | x | x | x | x | x |

**S10**- CERQual Assesment

| | | | | | | | | | | |
|---|---|---|---|---|---|---|---|---|---|---|
| **Water cost** | **Domestic** | Barrier | Mohamed 2022<br>Yallew 2012 | 4 | Minor concerns | Moderate concerns | Serious concerns | Serious concerns | Very Low | Study quality concerning; number of studies limiting adequacy and geographic relevance (only Africa). |
| | | Enabler | x | x | x | x | x | x | x | x |
| | **Institutional** | Barrier | x | x | x | x | x | x | x | x |
| | | Enabler | x | x | x | x | x | x | x | x |
| | **Public** | Barrier | x | x | x | x | x | x | x | x |
| | | Enabler | x | x | x | x | x | x | x | x |
| **Household infra- structure** | **Domestic** | Barrier | Atuyambe 2011<br>Biswas 2017<br>Didier 2021<br>Hoque 2023<br>Lohiniva 2007 | 5 | No concerns | Minor concerns | Minor concerns | Minor concerns | High | Overall, adequate number of studies supporting theme across multiple settings. Limitation in geographic relevance (mostly rural). |
| | | Enabler | Biswas 2017 | 5 | No concerns | Moderate concerns | Serious concerns | Serious concerns | Very Low | Limited number of studies limiting adequacy and geographical/population relevance. |
| | **Institutional** | Barrier | x | x | x | x | x | x | x | x |
| | | Enabler | x | x | x | x | x | x | x | x |
| | **Public** | Barrier | x | x | x | x | x | x | x | x |
| | | Enabler | x | x | x | x | x | x | x | x |

**S10**- CERQual Assesment

| | | | | | | | | | | |
|---|---|---|---|---|---|---|---|---|---|---|
| Community infra- structure | Domestic | Barrier | x | x | x | x | x | x | x | x |
| | | Enabler | x | x | x | x | x | x | x | x |
| | Institutional | Barrier | La Con 2017<br>Green 2005<br>Kumar 2018<br>Scott 2007<br>Sebong 2021<br>Steiner-Asiedu 2011 | 5 | No concerns | Minor concerns | Minor concerns | Minor concerns | High | Overall, adequate number of studies supporting theme across multiple settings. Limitation in geographic relevance (mostly LMICs, mostly schools). |
| | | Enabler | Jackson 2021 | 5 | No concerns | Moderate concerns | Serious concerns | Serious concerns | Very Low | Number of studies limiting adequacy and geographical/population relevance (USA only, athlectic raining facility only. |
| | Public | Barrier | Aberese-Ako 2023<br>Scott 2007<br>Yardley 2011 | 5 | No concerns | Minor concerns | Serious concerns | Serious concerns | Low | Number of studies limiting adequacy and geographical/population relevance. |
| | | Enabler | x | x | x | x | x | x | x | x |
| Hand hygiene station location | Domestic | Barrier | Akter 2022<br>Chidziwisano 2019<br>Hoque 2023<br>Nizame 2013<br>Ashraf 2017<br>Steiner-Asiedu 2011 | 5 | No concerns | Minor concerns | Minor concerns | Minor concerns | High | Overall, adequate number of studies supporting theme across multiple settings. Limitation in geographic relevance (mostly Asia, all rural). |
| | | Enabler | Biswas 2017<br>Ashraf 2017<br>Tibbels 2022 | 5 | No concerns | Minor concerns | Serious concerns | Serious concerns | Low | Limitation in geographic relevance (LMIC only). |
| | Institutional | Barrier | Al-Naggar 2013<br>La Con 2017<br>Green 2005<br>Jackson 2021 | 5 | No concerns | Minor concerns | Moderate concerns | Moderate concerns | Moderate | Limitation in geographic relevance (mostly edcaiton settings). |
| | | Enabler | Jackson 2021<br>Okello 2019<br>Pragle 2007 | 5 | No concerns | Minor concerns | Serious concerns | Serious concerns | Low | Limitation in geographic relevance (LMIC and HIC but limited number; schools and workplace, but limited). |

**S10**- CERQual Assesment

| | | | | | | | | | | |
|---|---|---|---|---|---|---|---|---|---|---|
| **Hand hygiene station location** | **Public** | Barrier | Watson 2020 | 5 | No concerns | Minor concerns | Serious concerns | Serious concerns | Very Low | Number of studies limiting adequacy and geographical/population relevance (IDP only) |
| | | Enabler | Neetu 2013 | 5 | No concerns | Minor concerns | Serious concerns | Serious concerns | Very Low | Number of studies limiting adequacy and geographical/population relevance (market only). |
| **Hand hygiene station design** | **Domestic** | Barrier | Hoque 2023 Mohamed 2022 White 2022 B Zangana 2020 | 4.3 | Minor concerns | Minor concerns | Moderate concerns | Moderate concerns | Moderate | Limitation in geographic relevance (mostly Africa; LMICs only). |
| | | Enabler | White 2022 A White 2022 B | 4.5 | Very minor concerns | Moderate concerns | Serious concerns | Serious concerns | Very Low | Number of studies limiting cogerence, adequacy and geographical/population relevance. |
| | **Institutional** | Barrier | x | x | x | x | x | x | x | x |
| | | Enabler | x | x | x | x | x | x | x | x |
| | **Public** | Barrier | Mohamed 2022 | 3 | Moderate concerns | Moderate concerns | Serious concerns | Serious concerns | Very Low | Low quality and number of studies limiting coerence, adequacy and geographical/population relevance. |
| | | Enabler | x | x | x | x | x | x | x | x |
| **State of hand hygiene equipment** | **Domestic** | Barrier | White 2020 B | 4 | Minor concerns | Moderate concerns | Serious concerns | Serious concerns | Very low | Limited number of studies limiting coherence, adequacy and geographical/population relevance. |
| | | Enabler | x | x | x | x | x | x | x | x |

**S10**- CERQual Assesment

| | | | | | | | | | | |
|---|---|---|---|---|---|---|---|---|---|---|
| **State of hand hygiene equipment** | **Institutional** | Barrier | Melaku 2023 | 5 | No concerns | Moderate concerns | Serious concerns | Serious concerns | Very low | Limited number of studies limiting coherence, adequacy and geographical/population relevance. |
| | | Enabler | Melaku 2023 | 5 | No concerns | Moderate concerns | Serious concerns | Serious concerns | Very low | Limited number of studies limiting coherence, adequacy and geographical/population relevance. |
| | **Public** | Barrier | Blum 2019 | 5 | No concerns | Moderate concerns | Serious concerns | Serious concerns | Very low | Limited number of studies limiting coherence, adequacy and geographical/population relevance. |
| | | Enabler | x | x | x | x | x | x | x | x |
| **Cleanliness of hand washing environ- ment** | **Domestic** | Barrier | Zangana 2020 | 5 | No concerns | Moderate concern | Serious concerns | Serious concerns | Very low | Limited number of studies limiting coherence, adequacy and geographical/population relevance. |
| | | Enabler | x | x | x | x | x | x | x | x |
| | **Institutional** | Barrier | Kumar 2018 | 5 | No concerns | Moderate concern | Serious concerns | Serious concerns | Very low | Limited number of studies limiting coherence, adequacy and geographical/population relevance. |
| | | Enabler | x | x | x | x | x | x | x | x |
| | **Public** | Barrier | Wu 2019 | 5 | No concerns | Moderate concern | Serious concerns | Serious concerns | Very low | Limited number of studies limiting coherence, adequacy and geographical/population relevance. |
| | | Enabler | x | x | x | x | x | x | x | x |

**S10**- CERQual Assesment

| | | | | | | | | | | |
|---|---|---|---|---|---|---|---|---|---|---|
| **Lighting of hand washing area** | **Domestic** | Barrier | x | x | x | x | x | x | x | x |
| | | Enabler | x | x | x | x | x | x | x | x |
| | **Institutional** | Barrier | x | x | x | x | x | x | x | x |
| | | Enabler | x | x | x | x | x | x | x | x |
| | **Public** | Barrier | Watson 2020 | 5 | No concerns | Moderate concern | Serious concerns | Serious concerns | Very low | Limited number of studies limiting coherence, adequacy and geographical/population relevance. |
| | | Enabler | x | x | x | x | x | x | x | x |
| **Damage of resources** | **Domestic** | Barrier | White 2022 A | 5 | No concerns | Moderate concern | Serious concerns | Serious concerns | Very low | Limited number of studies limiting coherence, adequacy and geographical/population relevance. |
| | | Enabler | x | x | x | x | x | x | x | x |
| | **Institutional** | Barrier | La Con 2017<br>Melaku 2023<br>Pragle 2007 | 5 | No concerns | Moderate concern | Serious concerns | Serious concerns | Low | Limited number of studies limiting coherence, adequacy and geographical/population relevance. |
| | | Enabler | x | x | x | x | x | x | x | x |
| | **Public** | Barrier | x | x | x | x | x | x | x | x |
| | | Enabler | x | x | x | x | x | x | x | x |
| **Theft of resources** | **Domestic** | Barrier | Akter 2014<br>Biswas 2017<br>Ntakirutimana 2021<br>Sultana 2018 | 5 | No concerns | Moderate concern | Moderate concern | Serious concerns | Low | Limited number of studies limiting coherence, adequacy and geographical/population relevance (3 of 4 in Bangladesh). |
| | | Enabler | x | x | x | x | x | x | x | x |
| | **Institutional** | Barrier | x | x | x | x | x | x | x | x |
| | | Enabler | x | x | x | x | x | x | x | x |
| | **Public** | Barrier | x | x | x | x | x | x | x | x |
| | | Enabler | x | x | x | x | x | x | x | x |

**S10**- CERQual Assesment

| | | | | | | | | | | |
|---|---|---|---|---|---|---|---|---|---|---|
| **Resources available for purchase** | **Domestic** | Barrier | Yeasmin 2021<br>Torres-Slimming 2019 | 4 | Minor concerns | Moderate concern | Serious concerns | Serious concerns | Very low | Limited number of studies limiting coherence, adequacy and geographical/population relevance |
| | | Enabler | x | x | x | x | x | x | x | x |
| | **Institutional** | Barrier | x | x | x | x | x | x | x | x |
| | | Enabler | x | x | x | x | x | x | x | x |
| | **Public** | Barrier | Thorseth 2021 | 5 | No concerns | Moderate concern | Serious concerns | Serious concerns | Very low | Limited number of studies limiting coherence, adequacy and geographical/population relevance |
| | | Enabler | x | x | x | x | x | x | x | x |
| **Social Opportunity** | | | | | | | | | | |
| **Role model for behavior** | **Domestic** | Barrier | Lanfer 2021 | 5 | No concerns | Moderate concerns | Serious concerns | Serious concerns | Very low | Number of studies limiting coherence, adequacy and geographical/population relevance |
| | | Enabler | Yeasmin 2021<br>Langford 2013<br>Sagan 2019<br>Ashraf 2017 | 4.3 | Minor concerns | Moderate concerns | Moderate concerns | Serious concerns | Low | Number of studies limiting coherence, adequacy and geographical/population relevance (all Bangladesh, Nepal, Pakistan) |
| | **Institutional** | Barrier | x | x | x | x | x | x | x | x |
| | | Enabler | x | x | x | x | x | x | x | x |
| | **Public** | Barrier | x | x | x | x | x | x | x | x |
| | | Enabler | x | x | x | x | x | x | x | x |

**S10**- CERQual Assesment

| | | | | | | | | | | |
|---|---|---|---|---|---|---|---|---|---|---|
| **Social Pressure** | **Domestic** | Barrier | Langford 2013 Parveen 2018 | 5 | No concerns | Moderate concerns | Serious concerns | Serious concerns | Very Low | Number of studies limiting coherence, adequacy, and geographical/population relevance |
| | | Enabler | Afolabi 2022 Harrison 2019 Langford 2013 Thaivalappil 2022 Tibbels 2022 | 5 | No concerns | Moderate concerns | Minor concerns | Moderate concerns | Moderate | Number of studies limiting coherence, adequacy, and geographical/population relevance |
| | **Institutional** | Barrier | x | x | x | x | x | x | x | x |
| | | Enabler | Green 2005 Mbakaya 2019 Pragle 2007 Schmidt 2009 | 5 | No concerns | Moderate concerns | Minor concerns | Moderate concerns | Moderate | Number of studies limiting coherence, adequacy, and geographical/population relevance (3 in HICs, 2 in Africa; School /workplace mix) |
| | **Public** | Barrier | x | x | x | x | x | x | x | x |
| | | Enabler | Blum 2019 | 5 | No concerns | Moderate concerns | Serious concerns | Serious concerns | Very low | Number of studies limiting adequacy and geographical/population relevance. |
| **Visits by Authorities** | **Domestic** | Barrier | x | x | x | x | x | x | x | x |
| | | Enabler | Ward 2022 | 5 | No concerns | Serious concerns | Serious concerns | Serious concerns | Very low | Number of studies limiting adequacy and geographical/population relevance Urban Nigeria only; COVID-related). |
| | **Institutional** | Barrier | x | x | x | x | x | x | x | x |
| | | Enabler | Sebong 2021 | 5 | No concerns | Serious concerns | Serious concerns | Serious concerns | Very low | Number of studies limiting adequacy and geographical/population relevance (univeraity in Asia only; COVID-related). |
| | **Public** | Barrier | x | x | x | x | x | x | x | x |
| | | Enabler | x | x | x | x | x | x | x | x |

**S10**- CERQual Assesment

| | | | | | | | | | | |
|---|---|---|---|---|---|---|---|---|---|---|
| **Social stigmatization/ status** | **Domestic** | Barrier | Azam 2022<br>Parveen 2018<br>White 2022 B<br>Yardley 2011 | 4.8 | Very minor concerns | Moderate concerns | Moderate concerns | Moderate concerns | Low | Number of studies limiting adequacy and geographical/population relevance. |
| | | Enabler | Langford 2013<br>Sagan 2019<br>Scott 2007<br>Tibbels 2022<br>White 2022 A<br>White 2022 B<br>Zangana 2020 | 4.9 | Very minor concerns | Minor concerns | Very minor concerns | Moderate concerns | Moderate | Number of studies limiting adequacy and geographical/population relevance (all LMICs). |
| | **Institutional** | Barrier | Kumar 2018 | 5 | No concerns | Moderate concerns | Serious concerns | Serious concerns | Very low | Number of studies limiting adequacy and geographical/population relevance. |
| | | Enabler | Green 2005<br>Jackson 2021 | 5 | No concerns | Moderate concerns | Serious concerns | Serious concerns | Very low | Number of studies limiting adequacy and geographical/population relevance. |
| | **Public** | Barrier | x | x | x | x | x | x | x | x |
| | | Enabler | Blum 2019<br>Watson 2020 | 5 | No concerns | Moderate concerns | Serious concerns | Serious concerns | Very low | Number of studies limiting adequacy and geographical/population relevance. |
| **Informational support** | **Domestic** | Barrier | Mohamed 2022 | 3 | Moderate concerns | Moderate concerns | Serious concerns | Serious concerns | Very low | Number of studies limiting adequacy and geographical/population relevance. |
| | | Enabler | Afolabi 2022<br>Torres-Slimming 2019 | 5 | No concerns | Moderate concerns | Serious concerns | Serious concerns | Very low | Number of studies limiting adequacy and geographical/population relevance. |
| | **Institutional** | Barrier | x | x | x | x | x | x | x | x |
| | | Enabler | x | x | x | x | x | x | x | x |
| | **Public** | Barrier | x | x | x | x | x | x | x | x |
| | | Enabler | x | x | x | x | x | x | x | x |

**S10**- CERQual Assesment

| | | | | | | | | | | |
|---|---|---|---|---|---|---|---|---|---|---|
| **Tangible/ instrumental support** | **Domestic** | Barrier | x | x | x | x | x | x | x | x |
| | | Enabler | Biswas 2017<br>Parveen 2018<br>Sultana 2018 | 5 | No concerns | Moderate concerns | Serious concerns | Serious concerns | Low | Number of studies limiting adequacy and geographical/population relevance (all Bangaldesh) |
| | **Institutional** | Barrier | x | x | x | x | x | x | x | x |
| | | Enabler | x | x | x | x | x | x | x | x |
| | **Public** | Barrier | x | x | x | x | x | x | x | x |
| | | Enabler | x | x | x | x | x | x | x | x |
| **Network support** | **Domestic** | Barrier | Rahman 2017<br>Parveen 2018<br>White 2022 A | 5 | No concerns | Moderate concern | Serious concerns | Serious concerns | Low | Number of studies limiting adequacy and geographical/population relevance. |
| | | Enabler | Dearden 2002<br>Langford 2013<br>Rahman 2017<br>Scott 2007<br>Parveen 2018<br>Thaivalappil 2022<br>White 2022 A | 5 | No concerns | Minor concern | Minor concern | Minor concern | Moderate | Limitid geographical relevance (mostly Africa, Asia). |
| | **Institutional** | Barrier | x | x | x | x | x | x | x | x |
| | | Enabler | Green 2005<br>Schmidt 2009 | 5 | No concerns | Moderate concern | Serious concerns | Serious concerns | Very low | Number of studies limiting adequacy and geographical/population relevance. |
| | **Public** | Barrier | x | x | x | x | x | x | x | x |
| | | Enabler | Watson 2020 | 5 | No concerns | Moderate concern | Serious concerns | Serious concerns | Very low | Number of studies limiting adequacy and geographical/population relevance. |

**S10**- CERQual Assesment

| | | | | | | | | | | |
|---|---|---|---|---|---|---|---|---|---|---|
| **Cultural practices and norms** | **Domestic** | Barrier | Aberese-Ako 2023<br>Hoque 2023<br>Kalumbi 2020<br>Parveen 2018<br>Ward 2022<br>White 2022 B | 4.8 | Very minor concerns | Minor concern | Minor concern | Moderate concern | Moderate | Limited geographical relevance (all Africa, Asia). |
| | | Enabler | Afolabi 2022<br>Chidziwisano 2019<br>Thaivalappil 2022<br>Zangana 2020 | 5 | No concerns | Moderate concern | Moderate concern | Serious concerns | Low | Limited geographical relevance (no Asia). |
| | **Institutional** | Barrier | x | x | x | x | x | x | x | x |
| | | Enabler | Jackson 2021 | 5 | No concerns | Serious concern | Serious concerns | Serious concerns | Very low | Limited number of studies limiting adequacy and geographical/population relevance. |
| | **Public** | Barrier | x | x | x | x | x | x | x | x |
| | | Enabler | x | x | x | x | x | x | x | x |

**S10**- CERQual Assesment

| MOTIVATION | | | | | | | | | |
|---|---|---|---|---|---|---|---|---|---|
| **Reflective Motivation** | | | | | | | | | |
| **Perceived health risk** | **Domestic** | Barrier | Azam 2022<br>Parveen 2018<br>Tibbels 2022<br>Thaivalappil 2022 | 5 | No concerns | Moderate concerns | Moderate concerns | Serious concerns | Low | Number of studies limiting adequacy and geographical/population relevance. |
| | | Enabler | Aberese-Ako 2023<br>Affleck 2012<br>Afolabi 2022<br>Akter 2014<br>Curtis 2003<br>Dearden 2002<br>Biswas 2017<br>Didier 2021<br>Kalumbi 2020<br>Lanfer 2021<br>Langford 2013<br>Lohiniva 2007<br>Nizame 2013<br>Norrie 2022<br>Ogutu 2022<br>Rahman 2017<br>Scott 2007<br>Parveen 2018<br>Steiner-Asiedu 2011<br>Tibbels 2022<br>Torres-Slimming 2019<br>Ward 2022<br>White 2022 B<br>Zangana 2020 | 5 | No concerns | No concerns | No concerns | Minor cocnerns | High | Adequate number of studies supporting theme across multiple settings |

**S10**- CERQual Assesment

| | | | | | | | | | | |
|---|---|---|---|---|---|---|---|---|---|---|
| **Perceived health risk** | **Institutional** | Barrier | Al-Naggar 2013<br>Sebong 2021 | 5 | No concerns | Serious concerns | Serious concerns | Serious concerns | Very low | Number of studies limiting adequacy and geographical/population relevance (asia only; Universities only). |
| | | Enabler | Mbakaya 2019<br>Okello 2019<br>Pragle 2007<br>Schmidt 2009<br>Sebong 2021 | 5 | No concerns | Minor concerns | Minor concerns | Minor concerns | Moderate | Adequate number of studies supporting theme across multiple settings, though mostly schools in Asia/Africa. |
| | **Public** | Barrier | x | x | x | x | x | x | x | x |
| | | Enabler | Babalobi 2013<br>Blum 2019<br>Watson 2020 | 5 | No concerns | Moderate concerns | Serious concerns | Serious concerns | Very low | Number of studies limiting coherence, adequacy and geographical/population relevance (Africa/Asia only; IDP camps and market). |
| **Time Prioritiza-tion** | **Domestic** | Barrier | Affleck 2012<br>Akter 2014<br>Curtis 2003<br>Dearden 2002<br>Langford 2013<br>Lohiniva 2007<br>Nizame 2016<br>Rauyajin 1994<br>Sagan 2019<br>Parveen 2018<br>Steiner-Asiedu 2011 | 5 | No concerns | No concerns | No concerns | Minor concerns | High | Overall, adequate number of studies supporting theme across multiple settings. Limitation in geographic relevance (mostly Asia, specifically Bangladesh). |
| | | Enabler | Dearden 2002<br>Biswas 2017 | 5 | No concerns | Serious concerns | Serious concerns | Serious concerns | Very low | Number of studies limiting adequacy and geographical/population relevance. |

**S10**- CERQual Assesment

| | | | | | | | | | | |
|---|---|---|---|---|---|---|---|---|---|---|
| | **Institutional** | Barrier | Al-Naggar 2013<br>La Con 2017<br>Green 2005<br>Jackson 2021<br>Pragle 2007<br>Schmidt 2009<br>Steenkamp 2022 | 5 | No concerns | No concerns | No concerns | Minor concerns | High | Overall, adequate number of studies supporting theme across multiple settings. Limitation in geographic relevance though mltiple regions and settings represented (though sometimes only once). |
| | | Enabler | x | x | x | x | x | x | x | x |
| | **Public** | Barrier | Blum 2019<br>Mezaache 2021<br>Neetu 2013<br>Nizame 2019<br>Wu 2019 | 4.4 | Minor concerns | Minor concerns | Minor concerns | Minor concerns | Moderate | Limitation in geographic relevance (mostly Asia, Africa); though multiple settings represented (e.g., Markets, parks, harm reduction centers).). |
| | | Enabler | x | x | x | x | x | x | x | x |
| **Water Prioritiza- tion** | **Domestic** | Barrier | Lohiniva 2007<br>Mshida 2020<br>Ogutu 2022<br>Thorseth 2021 | 5 | No concerns | No concerns | Moderate concerns | Moderate concerns | Moderate | Limitation in geographic relevance (all rural Africa). |
| | | Enabler | x | x | x | x | x | x | x | x |
| | **Institutional** | Barrier | x | x | x | x | x | x | x | x |
| | | Enabler | x | x | x | x | x | x | x | x |
| | **Public** | Barrier | x | x | x | x | x | x | x | x |
| | | Enabler | x | x | x | x | x | x | x | x |

**S10**- CERQual Assesment

| | | | | | | | | | | |
|---|---|---|---|---|---|---|---|---|---|---|
| **Willingness to practice hand-washing** | **Domestic** | Barrier | Affleck 2012<br>Didier 2021<br>Herbst 2009<br>Greenwell 2013<br>Mohamed 2022<br>Rauyajin 1994<br>Parveen 2018 | 4.7 | Very minor concerns | Moderate concerns | Moderate concerns | Minor concerns | Moderate | Overall, adequate number of studies supporting theme across multiple settings, though variability within theme across settings limiting coherence. |
| | | Enabler | Akter 2014<br>Dearden 2002<br>Harrison 2019<br>Herbst 2009<br>Rahman 2017<br>Parveen 2018 | 5 | No concerns | Moderate concerns | Moderate concerns | Minor concerns | Moderate | Overall, adequate number of studies supporting theme across multiple settings, though variability within theme across settings limiting coherence. |
| | **Institutional** | Barrier | Al-Naggar 2013<br>Green 2005 | 5 | No concerns | Moderate concerns | Serious concerns | Serious concerns | Very low | Number of studies limiting adequacy and geographical/population relevance. |
| | | Enabler | Pragle 2007 | 5 | No concerns | Moderate concerns | Serious concerns | Serious concerns | Very low | Number of studies limiting adequacy and geographical/population relevance. |
| | **Public** | Barrier | Thorseth 2021 | 5 | No concerns | Moderate concerns | Serious concerns | Serious concerns | Very low | Number of studies limiting adequacy and geographical/population relevance. |
| | | Enabler | x | x | x | x | x | x | x | x |

**S10**- CERQual Assesment

| | | | | | | | | | | |
|---|---|---|---|---|---|---|---|---|---|---|
| **Perceived ease or difficulty of washing hands** | **Domestic** | Barrier | Dearden 2002<br>Norrie 2022<br>Ward 2022<br>White 2022 A<br>Yardley 2011 | 5 | No concerns | Moderate concerns | Moderate concerns | Minor concerns | Moderate | Overall, somewhat limited number of studies supporting theme across multiple settings, and variability within theme across settings limiting coherence. Number limiting adequancy and relevance. |
| | | Enabler | Biswas 2017<br>Langford 2013<br>Mohamed 2022<br>Simiyu 2020<br>Sultana 2018<br>Ward 2022<br>White 2022 A | 4.7 | Very minor concerns | Moderate concerns | Minor concerns | Moderate concerns | Moderate | Overall, variability within theme across settings limiting coherence. Relevance limiting (All Africal, Asia). |
| | **Institutional** | Barrier | Steenkamp 2022 | 5 | No concerns | Moderate concerns | Serious concerns | Serious concerns | Very low | Number of studies limiting coherence, adequacy, and geographical/population relevance. |
| | | Enabler | Randle 2013 | 5 | No concerns | Moderate concerns | Serious concerns | Serious concerns | Very low | Number of studies limiting coherence, adequacy, and geographical/population relevance. |
| | **Public** | Barrier | Mezaache 2021<br>Nizame 2019<br>Wu 2019 | 4 | Minor concerns | Moderate concerns | Serious concerns | Serious concerns | Very low | Number of studies limiting coherence, adequacy, and geographical/population relevance. |
| | | Enabler | Mezaache 2021 | 2 | Serious concerns | Moderate concerns | Serious concerns | Serious concerns | Very low | Number of studies limiting coherence, adequacy, and geographical/population relevance. |

**S10**- CERQual Assesment

| | | | | | | | | | | |
|---|---|---|---|---|---|---|---|---|---|---|
| **Influence of religion** | **Domestic** | Barrier | Azam 2022<br>Sagan 2019<br>Tibbels 2022 | 5 | No concerns | Moderate concerns | Serious concerns | Serious concerns | Very low | Number of studies limiting coherence, adequacy, and geographical/population relevance. |
| | | Enabler | Afolabi 2022 | 5 | No concerns | Serious concerns | Serious concerns | Serious concerns | Very low | Number of studies limiting coherence, adequacy, and geographical/population relevance. |
| | **Institutional** | Barrier | x | x | x | x | x | x | x | x |
| | | Enabler | x | x | x | x | x | x | x | x |
| | **Public** | Barrier | x | x | x | x | x | x | x | x |
| | | Enabler | x | x | x | x | x | x | x | x |

**S10**- CERQual Assesment

| **Automatic Motivation** | | | | | | | | | |
|---|---|---|---|---|---|---|---|---|---|
| **Internal motivation/ habit** | Domestic | Barrier | Aberese-Ako 2023<br>Affleck 2012<br>Akter 2014<br>Atuyambe 2011<br>Azam 2022<br>Curtis 2003<br>Dearden 2002<br>Didier 2021<br>Kalumbi 2020<br>Langford 2013<br>Lohiniva 2007<br>Nizame 2013<br>Nizame 2016<br>Rauyajin 1994<br>Parveen 2018<br>Thaivalappil 2022<br>Tibbels 2022 | 5 | No concerns | Minor concern | No concern | Minor concern | High | Overall, adequate number of studies supporting theme across multiple settings. |
| | | Enabler | Affleck 2012<br>Bauza 2021<br>Biran 2005<br>Chidziwisano 2019<br>Curtis 2003<br>Dearden 2002<br>Biswas 2017<br>Didier 2021<br>Greenwell 2013<br>Kalumbi 2020<br>Lando 2018<br>Langford 2013<br>Mitchell 2021<br>Sagan 2019<br>Sedekia 2022<br>Parveen 2018<br>Sultana 2018<br>Thaivalappil 2022<br>Yardley 2011 | 5 | No concerns | No concerns | No concerns | Minor concerns | High | Overall, adequate number of studies supporting theme across multiple settings. |

**S10**- CERQual Assesment

| | | | | | | | | | | |
|---|---|---|---|---|---|---|---|---|---|---|
| **Internal motivation/ habit** | Institutional | Barrier | Al-Naggar 2013 Kumar 2018 Schmidt 2009 | 5 | No concerns | Moderate concners | Serious concerns | Serious concerns | Low | Number of studies limiting adequacy and geographical/population relevance (mostley educaitonal settings) |
| | | Enabler | Arendt 2015 Devkota 2020 Jackson 2021 Neetu 2013 Okello 2019 Pragle 2007 | 5 | No concerns | Moderate concners | Moderate concners | Moderate concners | Moderate | Theme represented cohrently and adequately though geographical/population relevance limited. |
| | Public | Barrier | Blum 2019 Mezaache 2021 Thorseth 2021 Wu 2019 | 4.2 | Minor concerns | Moderate concners | Moderate concners | Moderate concners | Low | Number of studies limiting adequacy and geographical/population relevance |
| | | Enabler | x | x | x | x | x | x | x | x |
| **Feeling of cleanliness** | **Domestic** | Barrier | x | x | x | x | x | x | x | x |
| | | Enabler | Afolabi 2022 Biran 2005 Curtis 2003 Demberere 2016 Ogutu 2022 Scott 2007 Parveen 2018 White 2022 A Zangana 2020 | 5 | No concerns | Minor concerns | Minor concerns | Minor concerns | High | Overall, adequate number of studies supporting theme across multiple settings. |
| | **Institutional** | Barrier | Schmidt 2009 | 5 | No concerns | Serious concners | Serious concerns | Serious concerns | Very low | Number of studies limiting adequacy and geographical/population relevance (UK school only) |
| | | Enabler | Schmidt 2009 | 5 | No concerns | Serious concners | Serious concerns | Serious concerns | Very low | Number of studies limiting adequacy and geographical/population relevance (UK school only) |
| | **Public** | Barrier | x | x | x | x | x | x | x | x |

**S10**- CERQual Assesment

| | | | | | | | | | | |
|---|---|---|---|---|---|---|---|---|---|---|
| **Feeling of cleanliness** | | Enabler | Blum 2019<br>Mezaache 2021<br>Neetu 2013<br>Wu 2019 | 4.2 | Minor concerns | Serious concners | Serious concners | Moderate concerns | Low | Number of studies limiting adequacy and geographical/population relevance (varied countries and locations, but ideas not saturated) |
| **Like/ dislike of handwash product** | **Domestic** | Barrier | Dearden 2002<br>Ward 2022 | 5 | No concerns | Moderate concnern | Serious concerns | Serious concerns | Very low | Number of studies limiting adequacy and geographical/population relevance |
| | | Enabler | Biran 2005<br>Langford 2013 | 5 | No concerns | Serious concerns | Serious concerns | Serious concerns | Very low | Number of studies limiting adequacy and geographical/population relevance |
| | **Institutional** | Barrier | x | x | x | x | x | x | x | x |
| | | Enabler | Randle 2013<br>Schmidt 2009 | 5 | No concerns | Serious concerns | Serious concerns | Serious concerns | Very low | Number of studies limiting adequacy and geographical/population relevance |
| | **Public** | Barrier | Mezaache 2021 | 2 | Serious concerns | Serious concerns | Serious concerns | Serious concerns | Very low | Number of studies limiting adequacy and geographical/population relevance |
| | | Enabler | Blum 2019<br>Mezaache 2021<br>Wu 2019 | 4 | Minor concerns | Moderate concerns | Moderate concerns | Serious concerns | Low | Number of studies limiting adequacy and geographical/population relevance |

*** No or very minor concerns / minor concerns / moderate concerns / serious concerns**

**** Is this synthesis finding "a reasonable representation of the phenomenon of interest?". Score categories: high, moderate, low, very low**
